# Partisan differences in health behaviors can impact respiratory disease dynamics

**DOI:** 10.64898/2026.01.14.26344076

**Authors:** Chris Soria, Audrey M. Dorélien, Dennis M. Feehan, Ayesha S. Mahmud

## Abstract

The transmission of respiratory pathogens is fundamentally shaped by human behaviors such as interpersonal contacts, use of face masks, and vaccination. Political party affiliation has been shown to be associated with health-related behaviors. Yet, partisan heterogeneity in health-related behaviors is typically not included in infectious disease transmission models. Here, we leveraged uniquely detailed data from the Berkeley Interpersonal Contacts Study (BICS) on partisan differences in contact rates, mask usage, and vaccination patterns during the first year of the COVID-19 pandemic. We find substantial differences in health-related behaviors by political affiliation. Republicans reported a significantly greater number of average daily contacts, lower propensity of using masks and of getting vaccinated for COVID-19. These findings hold even after controlling for observable demographic and location-based differences across survey respondents. We adapt the classic Susceptible-Infected-Recovered (SIR) model to incorporate partisan-specific behaviors and varying levels of political homophily to simulate an outbreak of a hypothetical respiratory pathogen. We find that the observed behavior differences lead to simulated Republicans experiencing higher infection and mortality rates and earlier peaks compared to Democrats. Incorporating greater within-group mixing further amplified partisan differences in disease outcomes. Finally, we show that failure to incorporate partisan behavioral heterogeneity in disease models can lead to inaccurate predictions about the size and timing of outbreaks in a population.

**Significance:** The timing and size of infectious disease outbreaks are shaped by health-related behaviors that affect disease transmission. Using data specifically designed to measure interpersonal contacts and other health behaviors, we find that during the COVID-19 pandemic, Republicans reported a significantly greater number of average daily contacts, lower propensity of using masks and of getting vaccinated. These observed differences lead to Republicans experiencing higher infection and mortality rates and earlier peaks compared to Democrats in a model simulating a hypothetical respiratory infection. Incorporating a preference for within-group mixing further amplified partisan differences in disease outcomes. Failure to incorporate partisan behavioral heterogeneity in disease models can lead to inaccurate predictions about the size and timing of outbreaks in a population.

## 1 Main

The dynamics of directly-transmitted respiratory diseases depend on human behaviors such as the daily number of interpersonal contacts and face mask use. Previous work has uncovered substantial demographic heterogeneities in these health-related behaviors (Adimora and Schoenbach, 2005; Okten, Gollwitzer and Oettingen, 2020); for example, age-specific differences in contact rates are a key driver of respiratory disease dynamics and age is therefore routinely incorporated in disease models (Schenzle, 1984; Ferguson, Nokes and Anderson, 1996; Dorélien et al., 2023; Feehan and Mahmud, 2021).

Partisan differences in mask-wearing, physical distancing, and vaccination emerged as salient features of the COVID-19 pandemic (Fan, Orhun and Turjeman, 2023; Gadarian, Goodman and Pepinsky, 2021, 2024), with differences often rivaling or exceeding those observed across age groups (Gollwitzer et al., 2020). These pandemic-era gaps continued a long-term trend of widening partisan differences in trust toward science and public health institutions (Blank and Shaw, 2015; Furnas, LaPira and Wang, 2025; Camacho-García et al., 2025) and in differential adherence to public health recommendations (de Hoog and Pat-El, 2024; Fraser et al., 2022). The persistence and breadth of these partisan divides suggest that partisan behavioral heterogeneity will likely persist in future disease outbreaks (Iyengar and Krupenkin, 2018; Heltzel and Laurin, 2020). But the epidemiological consequences of partisanship remain largely unquantified and partisanship is not typically included in disease models (Pacheco et al., 2024; Bedson et al., 2021).

Here, we measure partisan differences in important health related behaviors and demonstrate the importance of incorporating partisan heterogeneities in the analysis of infectious disease dynamics. We make three main contributions. First, we leverage uniquely detailed survey data on contact rates, partisanship, and behavior collected during the first year of the COVID-19 pandemic to characterize partisan differences in health-related behaviors and to identify their correlates. Second, we incorporate these empirically estimated partisan differences into widely used mathematical transmission models that simulate the spread of respiratory infectious diseases in a population. Third, we use these simulations to demonstrate the impact of partisan behavioral differences on both aggregate epidemic dynamics and partisan-specific health outcomes and disparities, following the introduction of a hypothetical novel pathogen.

We find that partisan differences in contact rates and mask usage are nearly as large as age-based differences and larger than racial and gender differences. On average, Republicans in our survey have higher contact rates, lower rates of mask usage, and lower rates of vaccination compared to Democrats. These disparities do not appear to be driven by place-based factors such as local mask mandates. Simulation results show that behavioral differences alone lead to Republicans experiencing higher infection and mortality rates compared to Democrats, as well as earlier peaks in disease incidence. These disparities are further amplified by increasing levels of within-group mixing and having a higher proportion of Republicans in the population. Lastly, we find that ignoring partisan differences in health behavior leads to inaccurate predictions of the timing and magnitude of disease peaks in the broader population. That is, models that exclude partisan heterogeneity in health behavior produce markedly different epidemic projections compared to models that incorporate it—even when both are based on the same population-average estimates of health behaviors.

## 2 Results

### 2.1 Are partisan differences in health-related behaviors large?

For partisanship to be worth incorporating as a dimension of behavioral heterogeneity in disease models, the behavioral differences it captures must satisfy two conditions: they must involve behaviors that directly affect transmission, and they must be large enough—relative to dimensions already incorporated in models—to plausibly matter for epidemic dynamics. We assess these conditions using data from the Berkeley Interpersonal Contacts Study (BICS), which collected direct measures of contact rates and mask usage—the key behavioral drivers of respiratory transmission—alongside partisan identity across the first 13 months of the COVID-19 pandemic in the United States. Estimates from BICS data revealed differences in contact rates, mask usage, and COVID-19 vaccination by political party (Table 1 and Table S1). Republicans reported an average of 6.6 daily contacts, while Independents reported 5.7 contacts and Democrats reported 5.3 contacts. Among those reporting contacts, 54.8% of Republicans wore masks, compared to 53.8% of Independents and 65.6% of Democrats. In Wave 6 (May 2021), self-reported vaccination rates were 57.1% for Republicans, 60.7% for Independents, and 74.0% for Democrats. These aggregate differences also varied over time during the pandemic (Table S2), with Republicans showing the highest volatility in contact patterns (SI Temporal Patterns), a pattern consistent with state-level analyses of partisan behavior change (Urmi et al., 2025).

**Table 1:** Pandemic Behaviors by Political Affiliation. Descriptive statistics from pooled BICS survey data (April 2020–May 2021) weighted for national representativeness. Contact behaviors measured as previous-day reports; mask use calculated among respondents reporting contacts; vaccination rates as of wave 6 (May 2021).

| Reported Behavior | Republicans | Independents | Democrats |
|---|---|---|---|
| Total Daily Contacts | 6.6 | 5.7 | 5.3 |
| Non-Household Contacts | 4.7 | 3.9 | 3.5 |
| % Contacts with a Mask | 54.8 | 53.8 | 65.6 |
| % Vaccinated | 57.1 | 60.7 | 74.0 |
| N | 5,517 | 4,527 | 8,189 |

Figure 1 illustrates the magnitude of these partisan differences in three health behaviors by comparing them with differences in the same behaviors across age, gender, race, and education groups. Republicans reported about 1.4 more average daily contacts than Democrats— a difference more substantial than the gap in average contact rates between genders, educational groups, or racial groups (Figure 1). Among those reporting contacts, Republicans and Independents were both 17% less likely than Democrats to report wearing masks (Figure 1). This partisan gap exceeded differences based on gender, race, and age. Age-structured contact patterns are incorporated in respiratory disease models precisely because age-based behavioral gaps are large enough to affect transmission dynamics (Schenzle, 1984; Dorélien et al., 2023); the observed partisan gaps in contact rates and mask usage are comparable in magnitude and, in the case of masking, larger. When restricting the analysis to respondents aged 65 and older, both the overall pattern and size of partisan differences remained very similar to the full sample (Fig. S2), indicating that age composition does not account for the observed partisan behavioral gaps.

**Figure 1.**
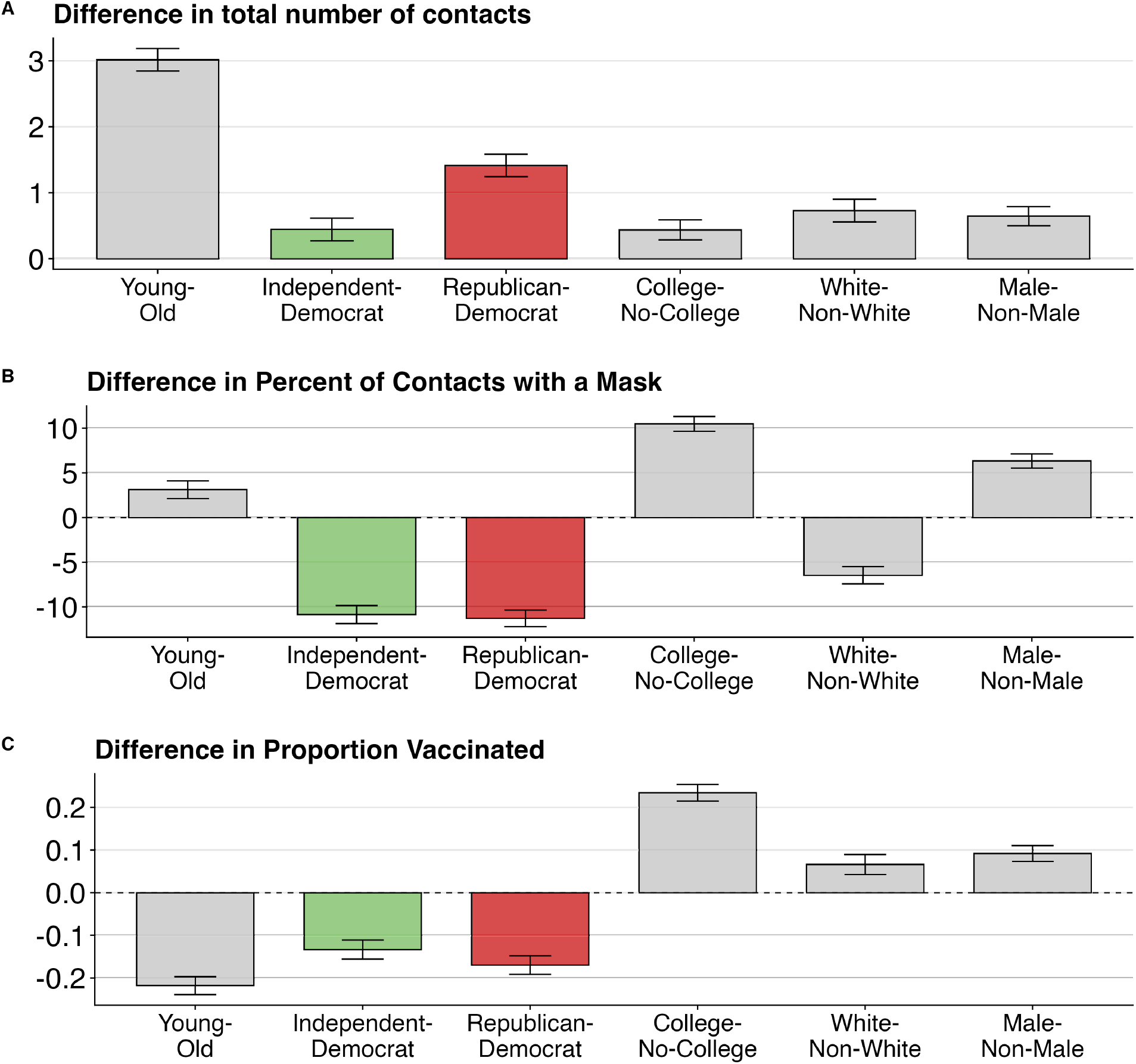
Comparison of partisan and demographic group differences for three health behaviors: (A) total reported in-person contacts from the previous day; (B) percentage of contacts involving mask use among those reporting previous-day contacts, based on detailed information from three selected contacts per respondent; and (C) estimated vaccination rates reported in Wave 6 of BICS in May 2021. Effect sizes are coefficients from univariate linear regression models estimated separately for each demographic characteristic and behavior. Sample includes respondents across six BICS survey waves (April 2020 – May 2021), except Panel C. Error bars show 95% confidence intervals.

Sociodemographic characteristics of BICS respondents differed across political parties: for example, Republicans in BICS were older, more likely to be white, and more likely to be male than Democrats (Table S3). These differences are consistent with other estimates of party composition (Zingher, 2018; Gadarian, Goodman and Pepinsky, 2021; Fan, Orhun and Turjeman, 2023). We use linear regression models to test whether partisan differences in each health behavior – contacts, masking, and vaccination – persist after controlling for other observed differences across individuals. In these regression models, we controlled for sociodemographic characteristics (race, gender, age group, ethnicity, education, employment, household size) and geographic-context covariates (county Democratic vote share, mortality rates, mask mandates, and state fixed effects).

Substantively large and statistically significant partisan differences persisted across all three health behaviors after controlling for individual and county-level characteristics, although estimated partisan differences were slightly attenuated (Figure 2). For example, after adding demographic, geographic, and policy controls, Republicans were predicted to have an average of 1.1 more contacts than Democrats (Table S4); Republicans exhibited 7.5 percentage-points lower mask use than Democrats (Table S5); and Republicans were roughly 16 percentage points less likely to be vaccinated than Democrats (Table S6).

**Figure 2.**
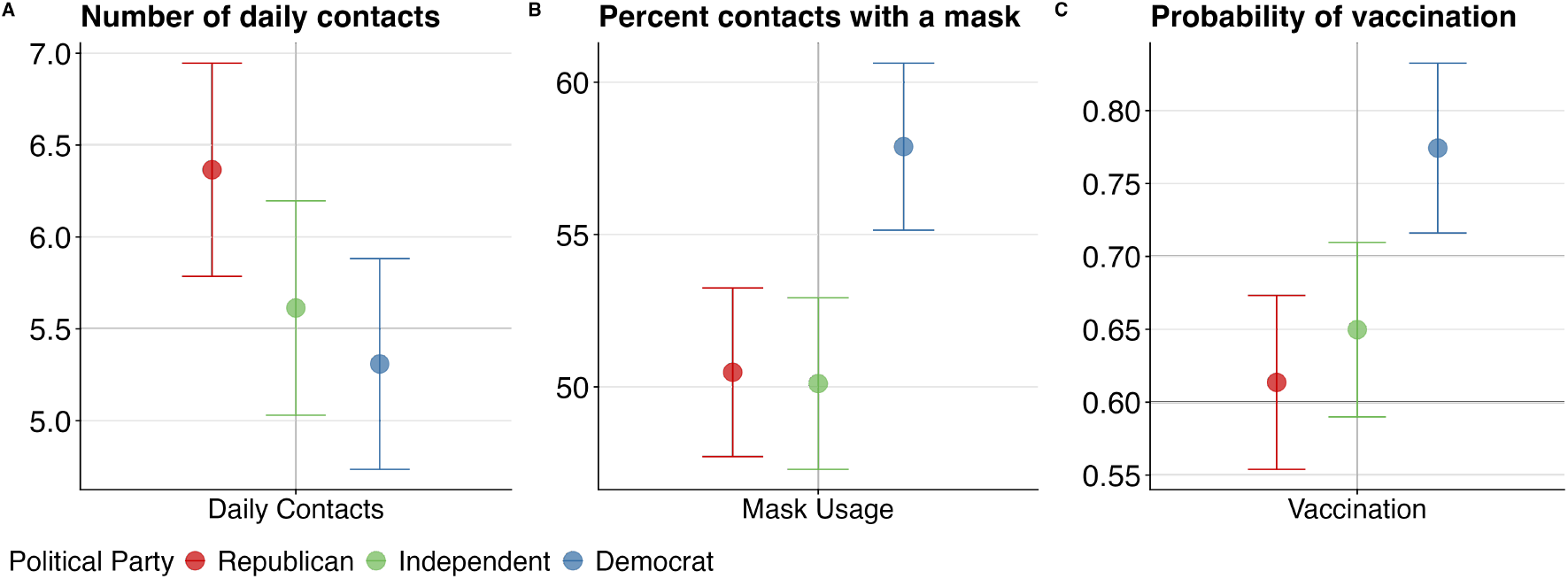
Adjusted effect sizes from multivariate linear regression models for (A) daily in-person contacts, (B) mask use, and (C) vaccination rates as of May 2021, controlling for survey design, demographics, and geographic context. Sample includes respondents across six BICS survey waves (April 2020 – May 2021), except Panel C which only includes respondents from Wave 6. Error bars represent 95% confidence intervals. The estimated marginal means are calculated for a reference individual who is a non-Hispanic White female aged 18-30, employed, college-educated, living in a metropolitan area in California during Wave 2 on a weekday, in a county with a strict mask mandate, with household size, log previous week mortality rate, and congressional district percent Democrat all set to their weighted sample averages.

Democrats also differed significantly from Independents in mask use and vaccination rates after adding controls, though these gaps were smaller than Republican-Democrat differences. These results indicate that the partisan association in behaviors directly relevant to transmission is not reducible to demographic composition, local policy context, or geographic sorting. The magnitude of this independent association rivals that of age—a dimension routinely incorporated in respiratory disease models. Partisanship is therefore a candidate dimension of behavioral heterogeneity that should be considered when modeling transmission in politically polarized populations.

### 2.2 Are partisan health-behaviors large enough to produce meaningful disease disparities?

To investigate the implications of these partisan differences for disease dynamics, we incorporated political heterogeneity in contact rates and protective behaviors into a mechanistic model for the transmission of a hypothetical respiratory pathogen. We adapted the classic Susceptible-Infected-Recovered (SIR) framework by subdividing the population into three distinct partisan groups, each with its own contact rate. To model the impact of mask-wearing, each partisan group is further divided into “protected” (P) and “unprotected” (U) subgroups (see Methods; Figure 6).

In our first set of results, we assume that the three political parties are equal in population size. We also assume that people mix randomly, i.e., without paying attention to political affiliation (this assumption is relaxed below). Remaining model parameters are chosen based on established values from the epidemiological literature (Table S7).

We evaluate three key outcomes of the mechanistic model: (1) cumulative incidence, which serves as a measure of total disease burden; (2) peak incidence, which summarizes the intensity of the outbreak; and (3) time to peak incidence, which serves as a measure of how rapidly the disease progresses through the population.

Figure 3 summarizes simulation results by comparing key disease outcomes across partisan groups, with all metrics standardized relative to Democrats (value = 1). Using only the aggregate behavioral differences in contact rates and mask usage documented above (Section 2.1)—while holding all other factors equal—simulated Republicans experience a 15% higher cumulative incidence than Democrats after one year. Republicans also face a peak incidence about 21% higher than Democrats, with the peak arriving roughly two days earlier. The higher transmission rate experienced by Republicans—due to higher contact rate and lower rate of adopting protective behavior—results in a quicker depletion of their susceptible pool and a larger proportion that are eventually infected and recovered, compared to Democrats and Independents (Fig. S4). In general, in our simulations, contact rates play a larger role than mask usage in driving these partisan differences in disease outcomes (SI Decomposing Effects).

**Figure 3.**
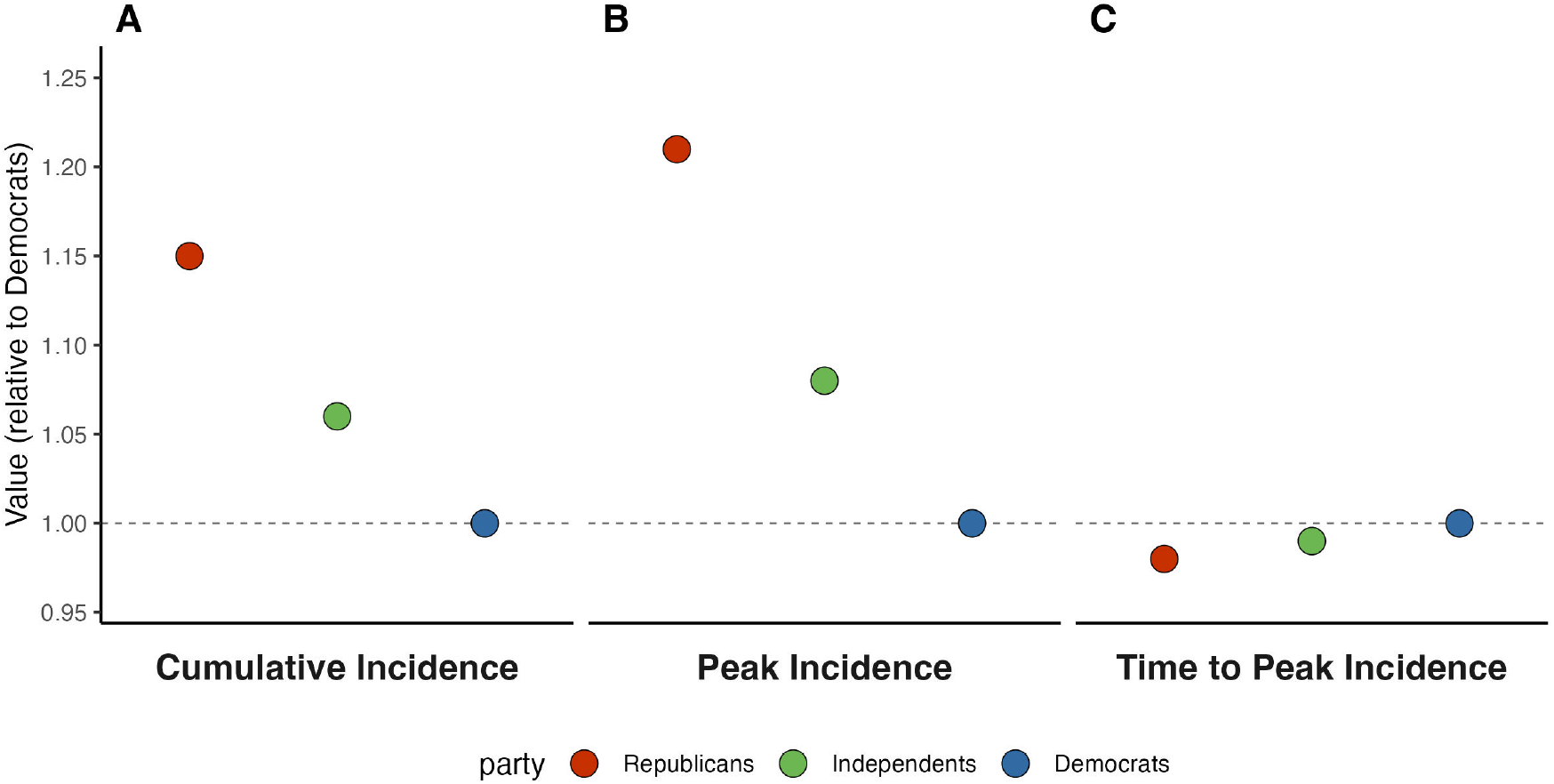
Simulation results from a three-party SIR model for (A) cumulative incidence (B) peak infection as a percentage of the population, and (C) time to peak infection. Results are from a one-year simulation for a hypothetical respiratory pathogen, with homogeneous partisan mixing and equal partisan group sizes. Partisan contact rates and rate of adoption of protective behavior are obtained from BICS data. All other model parameters were sourced from published literature as detailed in Table S5. All outcomes are shown as values relative to Democrats (reference = 1); > 1 indicates higher than Democrats, *<* 1 indicates lower than Democrats.

Many infectious diseases, such as COVID-19 and influenza, have a strong age-related mortality gradient (Sorci, Faivre and Morand, 2020); if the age composition of political parties differ, this could lead to different case fatality rates for each political party. As a sensitivity analysis, we allow the case fatality rate to vary across partisan groups to account for differences in their age composition; this produces a higher case fatality rate for Republicans, who tend to be older. In this scenario, the mortality results change dramatically: Republicans show a 75% higher mortality rate than Democrats (Fig. S6), a difference that reflects the combined effect of their older age distribution and behavioral factors.

To gauge how much partisan behavior drives respiratory infectious disease dynamics, we also reran the simulations under a counterfactual “cautious population” scenario. In this scenario, all partisan groups adopt the population-average behavior of the partisan group with the lowest-transmission profile observed in the survey, i.e. the fewest social contacts combined with the highest mask-use rate. Fig. S5 shows that moving the entire population to this best-case behavioral scenario would reduce the epidemic peak by 47% and avert about 18% of deaths relative to the simulations with partisan heterogeneity in behavior. All groups have lower incidence and mortality, but the benefits are unequally distributed: Democrats would experience 12% fewer deaths, while Republican deaths would drop by 25% and their peak caseload would fall by 51% (Table S8). In short, if every partisan group adopted the behaviors of the most cautious group, the outbreak’s overall severity would decline dramatically. A population universally adopting this lowest-transmission behavior would also markedly delay the time needed to reach the outbreak peak by nearly three months, extending the epidemic’s duration while reducing its intensity.

### 2.3 Do homophily and population composition moderate partisan disease disparities?

Having established that partisan behavioral differences alone produce meaningful disease disparities (Section 2.2), we next examined how partisan homophily and population composition moderate these outcomes. We accounted for the possibility of homophily by political affiliation in our model by including a parameter, *β*, that controls the extent to which contacts are disproportionately likely to happen within partisan groups. In the random mixing scenario discussed above, *β* = 1; when *β >* 1, contacts are more likely within partisan groups. We accounted for partisan composition in our model by including two parameters that govern the share of the population that is Republican and Democrat. We repeated the simulation described above for all combinations of *β* ∈ [1, 5] and share of Republicans and Democrats between 20 and 70%.

Figure 4 shows how homophily and population composition interact to influence cumulative infections after one year among Republicans (panel A) and Democrats (panel B). For instance, in Figure 4 Panel A, coordinates (1, 0.2) show that when *β* = 1 (random mixing) and when Republicans constitute 20% of the population, (and Democrats 70%, Independents 10%), about 95% of Republicans end up infected after a year. Conversely, at the opposite extreme where homophily is high (e.g., *β* = 5) and Republicans make up 70% of the population, the cumulative infections exceed 100% among Republicans (this includes repeat infections). Figure 4 reveals that as homophily increases (*β* increases along the x-axis) and a greater share of contacts occur within partisan groups, cumulative infections for Repbulicans increase and cumulative infections for Democrats decrease. As the Republican fraction of the population increases (increasing values along the y-axis), a greater number of contacts for all parties occurs with Republicans, increasing cumulative infections for both Republicans and Democrats (Fig. S7 shows the underlying partisan-specific epidemic curves for selected scenarios).

**Figure 4.**
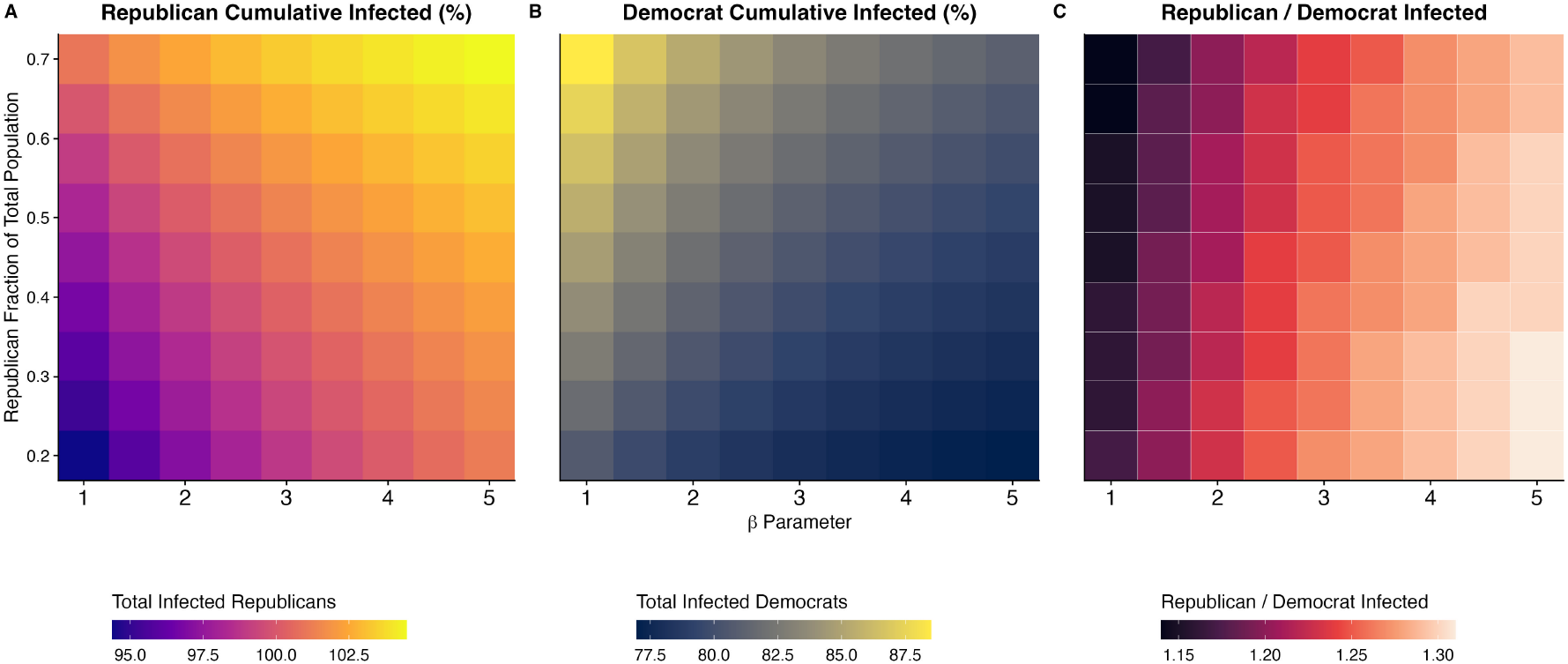
Simulated cumulative infections for (A) Republicans and (B) Democrats, and (C) the ratio of the number of Republican infections to Democrat infections. Results are from simulations with varying Republican homophily parameters (*β*: 1.0-5.0) and population fractions using the three-party SIR model with base parameters from Table S7. X-axis shows increasing partisan homophily (higher values = stronger in-group contact probability); y-axis shows Republican population proportion (20-70%) with Democrats comprising the remainder and Independents fixed at 10%.

Fig. S8 shows parallel results for the other disease outcomes. Republican mortality, peak incidence, and timing of peak infections become more severe with stronger partisan homophily and larger Republican population shares; both factors increase within-group contacts among Republicans (Fig. S11), who have higher contact rates and lower rates of adpoting protective behaviors. This increases the force of infection for Republicans, thereby increasing disease burden (Figure 4A; Fig. S8A,C,E). In scenarios combining high homophily and a large Republican majority, cumulative incidence surpasses 100% (Table S9), with epidemic peaks occurring up to 46 days earlier compared to Democrat-majority populations under random mixing. However, when Republicans represent 70% of the population, increasing homophily from *β* = 1 to *β* = 5 advances the Republican peak by only 5 days, indicating reduced sensitivity of peak timing to homophily under Republican-majority conditions (Fig. S8E-F).

Democrat cumulative incidence, on the other hand, decreases as both homophily and the proportion of Democrats in the population increase (Figure 4B). Higher homophily benefits Democrats by increasing within-group contacts and reducing their contacts with Republicans ((Fig. S12), thereby lowering their force of infection. Higher homophily also increases disparities between Democrats and Republicans’ cumulative incidence rates; the highest disparities occur when share of Democrats are high (see Figure 4C). Even in contexts where Democrats are a small minority within Republican-majority populations, increasing homophily consistently reduces Democrat cumulative incidence. However, as the Republican share of the population rises, Democrats experience increased cumulative incidence due to greater number of contacts with Republicans. Across all simulations with varying homophily and population composition, Democrat cumulative incidence rates (Fig. S9 B,D) consistently remain below Republican incidence rates (Fig. S9, A,C).

In the previous section, we show that when there is no homophily and partisan groups are equal in size, Republican cumulative incidence is 15% higher than that of Democrats. Introducing partisan homophily and variation in population composition greatly affects this relative difference in outcomes. For example, when Republicans constitute 50% of the population and exhibit high within-group mixing (*β* = 5), cumulative incidence is 31% higher in Republicans than in Democrats (Figure 4, Panel C).

Similarly, mortality, peak incidence, and timing of peak infections among Democrats generally decrease as homophily and the proportion of Democrats in the population increase (Figure 4B and Fig. S8B,D,F).

### 2.4 Do partisan behavioral differences impact overall epidemic projections?

The vast majority of respiratory disease transmission models used for simulations and forecasts either do not account for heterogeneities in the force of infection across population subgroups, or only do so by age.

We find large differences in contact rates and other behaviors observed across partisan groups, yet disease models do not typically incorporate political heterogeneity.

Here, we investigate whether or not ignoring partisan differences has an impact on population-level disease models in a setting where partisan differences are large. We construct a hypothetical population in which partisan health behaviors match average values observed in BICS, and assuming high levels of homophily (*β* = 5). We call this the *homophilous partisan heterogeneity model*. Using this model as the reference, we then investigate how well the aggregate dynamics of the outbreak can be approximated using two simpler models: (1) the *partisan heterogeneity model*, which assumes that contact and mask usage rates vary by party, but that does not account for homophily; and (2) the *uniform behavior model*, which disregards partisanship completely and instead models population average behaviors observed in BICS.

To explore how population composition moderates these effects, we examine each model across three distinct population compositions: an evenly split population (R: 33%, D:33%, I: 33%), a Republican-majority population (R: 70%, D: 20%, I: 10)%, and a Democrat-majority population (R: 20%, D: 70%, I: 10%).

Figure 5 shows the results for an evenly split population (panel A), for a heavily Republican population (panel B), and for a heavily Democratic population (panel C). Within each of these Panels A, B, and C in 5, we assume the same population-average contact rates and health behaviors for all three scenarios. In an evenly split population, the simpler models (Uniform Behavior and Partisan Heterogeneity) predict a similar cumulative incidence rate as the Homophilous Partisan Heterogeneity model but over-estimate the time it takes for the outbreak to reach its peak: the partisan heterogeneity model predicts peak incidence 8 days late, and the uniform behavior model predicts peak incidence 20 days late (panel A; Table S8, Table S10).

**Figure 5.**
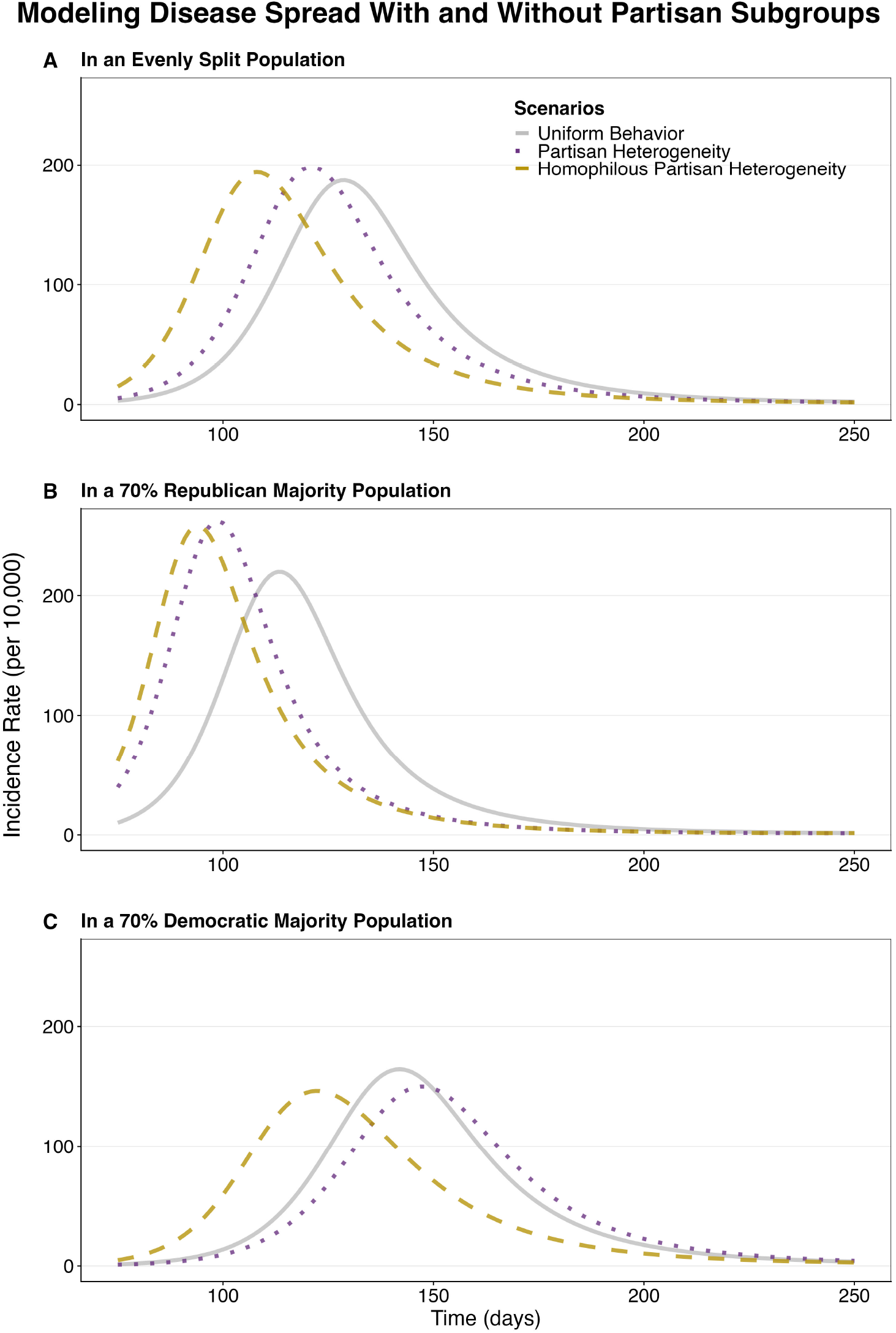
Simulated incidence rates per 10,00 in (A) an evenly split population, (B) a Republican majority population, and (C) a Democrat majority population. Gray solid line shows theuniform behavior model using population-average behaviors; purple dotted line shows the same population with partisan behavioral heterogeneity; yellow dashed line shows the partisan heterogeneity model with strong in-group preference (*β*=5).

**Figure 6.**
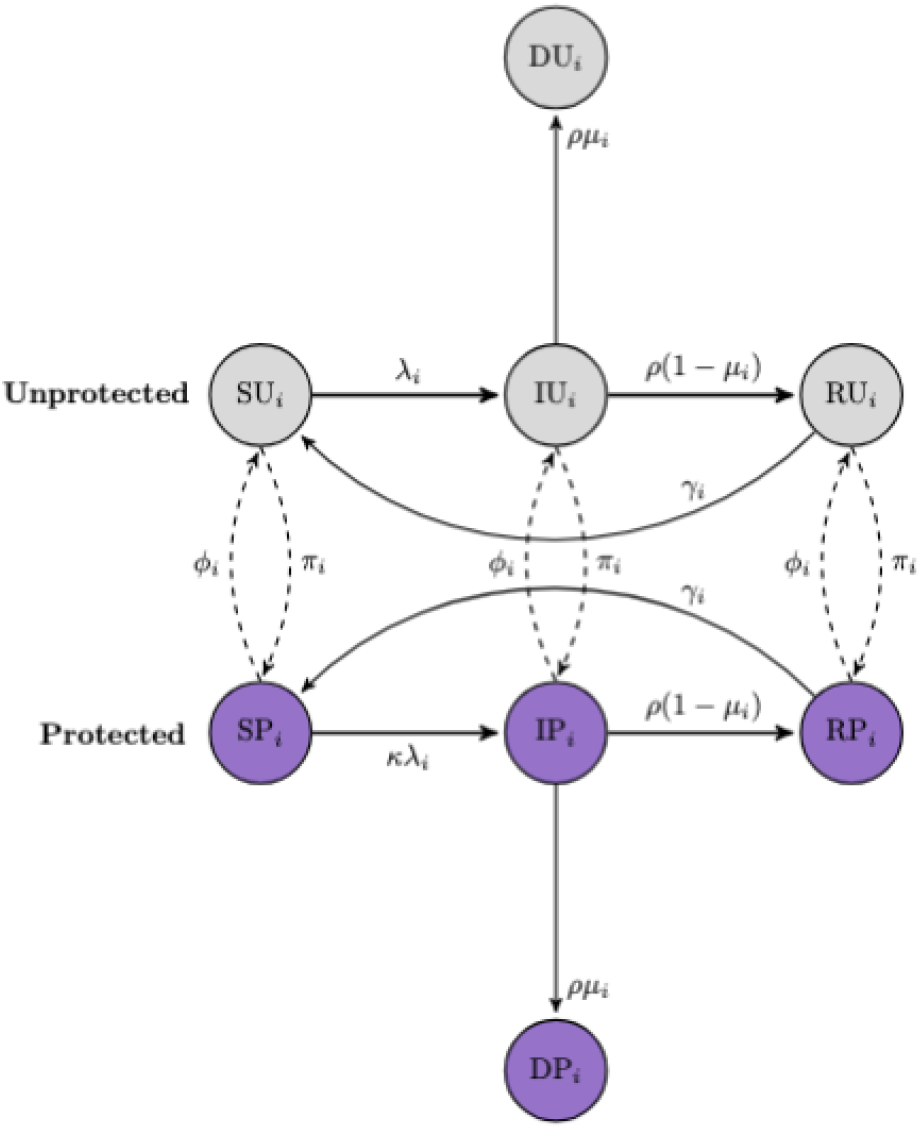
Flow diagram showing movement between unprotected and protected SIR classes

In a population where Republicans are the majority, both of the simpler models have a later peak compared to the Homophilous Partisan Heterogeneity scenario (Figure 5 B and Table S9). Relative to this reference, the Partisan Heterogeneity Model without homophily delays the peak by 4 days with marginally different outcomes. The Uniform Behavior Model shows the largest deviations: delaying the peak by 19 days, reducing peak prevalence from 2,100 to 1,869 per 10,000, and lowering cumulative mortality from 135 to 114 deaths per 10,000. In Republican-majority populations, homophily produces comparatively smaller changes in epidemic projections because demographic composition alone ensures high intragroup contact rates among Republicans, even under random mixing assumptions (Fig. S11, B).

In a population where Democrats are the majority, the Partisan Heterogeneity Model without homophily delays the peak by 24 days and moderately increases the final outbreak size compared to the Homophilious Partisan Heterogeneity model (Table S11). The Uniform Behavior Model shows even larger deviations, delaying the peak by 18 days from the reference while substantially increasing cumulative mortality. In this Democratic-majority composition, the minority Republican subgroup drives these epidemic dynamics, as homophily concentrates their within-group interactions (Fig. S11, C), accelerating overall transmission while reducing peak infections among the Democratic majority.

Overall, these results demonstrate that incorporating partisan heterogeneity and homophily into epidemiological models is critical for accurate outbreak projections, as simpler approaches systematically underestimate epidemic speed and, depending on population composition, may substantially misjudge disease burden.

## 3 Discussion

Our analysis produced three key insights: first, partisan disparities in health behaviors are substantial in magnitude even after controlling for demographic and geographic factors; second, incorporating these substantial partisan differences in disease models reveals corresponding disparities in outbreak timing and disease burden; and third, these partisan differences substantially alter aggregate disease spread projections, particularly when modeled with assumptions about partisan homophily.

Partisan differences in protective health behaviors are more pronounced than those associated with gender, race, or education—factors widely considered meaningful enough for inclusion in epidemiological models (Manna et al., 2024; Buckee, Noor and Sattenspiel, 2021; Tizzoni et al., 2022; Bedson et al., 2021). These partisan gaps cannot be attributed to demographic or geographic variation, both of which are commonly adjusted for when accounting for behavioral heterogeneity in disease modeling (Mistry et al., 2021).

Disease models reveal that these partisan behavioral differences translate into significant disparities in partisan-specific disease outcomes. Under random mixing contact patterns (*β* = 1) where partisan groups are assumed equal in size, Republicans experience 15% higher incidence after one year; when combined with their older age distribution, this results in 75% higher mortality (Figure 3). When we incorporate homophilous mixing, disparities increase substantially (up to 31% higher relative incidence), highlighting the importance of incorporating partisan homophily into disease models (Figure 4); this finding is consistent with other studies that have shown that homophily can substantially alter epidemic dynamics (Smaldino and Jones, 2021; Thomas et al., 2020). Research shows that levels of partisan homophily in contact are plausibly high (Kalmoe and Mason, 2022; Iyengar and Westwood, 2015; Iyengar and Krupenkin, 2018; Brown and Enos, 2021; Huber and Malhotra, 2017) but, unfortunately, no existing contact surveys directly measure partisan homophily. Since it seems unlikely that survey respondents can reliably identify their contacts’ political affiliations, future research should prioritize developing statistical methods that estimate partisan homophily indirectly through observable network and geographic patterns.

Partisan heterogeneity also fundamentally alters population-level epidemic projections by shifting outbreak timing and magnitude, even when simple models accurately capture overall population behaviors. Models ignoring partisan heterogeneity make inaccurate predictions about critical outcomes such as time to outbreak peak, with the largest discrepancies occurring in Democrat-majority populations (Figure 5). Although a one- to two-week discrepancy may seem modest, such mistiming could critically impair hospital surge planning, resource allocation, with potentially substantial mortality impacts (Geoffrey French et al., 2021; Amuedo-Dorantes, Kaushal and Muchow, 2021)

Several limitations of our analysis point to important directions for future research. First, we do not have data on partisan differences in contact rates *prior* to the COVID-19 pandemic. Although our analysis focuses on health behavior data collected during the COVID-19 pandemic, partisan differences in health behaviors are not new; longstanding and widening partisan divides in attitudes toward science (Kennedy, 2024), health policy (Fraser et al., 2022; Gollust et al., 2024), and vaccination (Baum, 2011; Jones and McDermott, 2022) suggest these behavioral gaps will most likely persist in future epidemic and pandemic contexts (Fan, Orhun and Turjeman, 2023; Gadarian, Goodman and Pepinsky, 2021, 2024). Future social contact studies are needed to determine whether these differences persist outside of pandemic contexts. Additionally, partisan health behaviors likely vary across geographic settings (Mistry et al., 2021), and future work should examine how local context shapes these behavioral patterns. Collecting data on prevalence- and mortality-dependent behavior by political party affiliation would also be valuable; although not shown in this paper, we found preliminary evidence that Republicans may exhibit mortality-dependent behavior, which could help reduce disparities in health outcomes. Finally, our analysis focused on health outcomes, but protective behaviors carry other costs and benefits—including economic, social, and psychological dimensions—that warrant further investigation. These data limitations are not merely technical gaps. Without accurate measurement of partisan homophily and the context-dependence of behavioral divides, models used to guide resource allocation, vaccination campaigns, and intervention timing may systematically misestimate outbreak risk in politically homogeneous communities—precisely the populations where behavioral clustering is strongest. Given that future pandemics are expected (Marani et al., 2021), these modeling errors could result in preventable loss of life.

## 4 Data and Methods

### Data

The Berkeley Interpersonal Contacts Study (BICS) used surveys to collect detailed data on demographic and socioeconomic characteristics, contact rates, and health-related behaviors during the first 13 months of the COVID-19 pandemic in the United States. Respondents were asked how many people they had face-to-face contact with on the day before the survey interview, and detailed information was collected on up to three of these contacts, including information about each contact’s age, sex, race/ethnicity, mask usage, and physical distance. Repeated cross sections were recruited from an online panel and weighting methods were developed to make the sample representative of the national population. Data collection started with a pilot wave in March 2020 (Wave 0) and continued with six additional waves conducted over the next year and a half. Additional details on the survey methodology, questionnaire design, sampling, and methods used for weighting are described in (Feehan and Mahmud, 2021).

We analyze data from BICS waves 1 through 6, spanning April 2020 to May 2021. Wave 0 is omitted because it did not record political variables in a comparable way. Political party identification was assessed by the panel provider prior to the survey. Upon enrollment in the panel, all potential survey respondents were asked to select their partisan affiliation from standard categories (Democrat, Republican, Independent, or Prefer not to Say/Other). We exclude respondents in the “Prefer not to Say/Other” category, to prevent potential mis-classification bias. The analytical sample therefore comprises respondents in three discrete self-identified partisan categories: Democrats, Republicans, and Independents.

We focus on three measures of health behavior. First, we examine **average daily contacts**, measured through the question “How many people did you have in-person contact with yesterday?” and through reports about the respondent’s household members. Second, for respondents reporting non-zero contacts outside the household, we assessed mask-wearing behavior for up to three contacts, measured through the question “During this contact, did you wear a face mask?” For this measure we calculate the **percent of contacts with a mask**. Third, we analyze self-reported COVID-19 vaccination status (“Have you gotten vaccinated against COVID-19?”) from Wave 6 (May 2021) data, when vaccines had become accessible to the general public. Our measure of interest is **percent vaccinated**.

### Statistical model to characterize partisan differences

We first investigate the relationship between partisan identity and health behaviors using regression models. We pool the data across survey waves and, for each of the three measures of health behavior (described above), we estimate the following regression model:

,
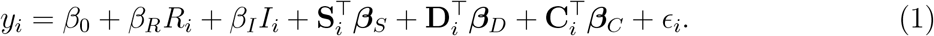

where *y*_*i*_ is the health-behavior measure for respondent *i*; *R*_*i*_ and *I*_*i*_ are indicators for Republican and Independent identification, respectively, with Democrats as the omitted category; **S**_*i*_ is a vector of survey characteristics (wave, day, city); **D**_*i*_ is a vector of sociodemographic characteristics for *i* (race, gender, age group, ethnicity, education, employment, household size); and **C**_*i*_ is a vector of geographic-context covariates (county Democratic vote share, mortality rates, mask mandates, and state fixed effects). The model uses survey weights to account for the study design.

The coefficients on the binary indicators for Republican (*β*_*R*_) and Independent (*β*_*I*_) represent the adjusted differences in health behaviors relative to Democrats, controlling for sociodemographic composition (see Table S3), survey characteristics, and geographic area.

We also estimated models more suited to the data structure (negative binomial for contact counts, logistic regression for vaccination outcomes), but results were substantively identical. We present linear models for ease of interpretation.

### Disease transmission model

We next investigate the implications of partisan differences in behavior for the dynamics of a hypothetical respiratory disease outbreak. We adapt an SIR model of respiratory disease transmission to capture heterogeneities in health-related behavior and interactions across the three partisan groups. In the SIR compartmental model, the population is divided into three mutually exclusive groups: susceptible, infected, and recovered. We also subdivide the population into three distinct partisan groups. To model the impact of mask-wearing, each partisan group is further divided into “protected” (P) and “unprotected” (U) subgroups (Harris, Cardenas and Mordecai, 2023; Smaldino and Jones, 2021) (Figure 6). Individuals in the protected subgroups have a lower probability than individuals in the unprotected subgroups of acquiring the infection following contact with an infected individual, as well as a lower probability of transmitting the infection to a susceptible individual. Unprotected individuals in political party *i* adopt protective behaviors at rate *π*_*i*_ and move to the P subgroup; to capture fatigue and waning of protective health behaviors, we model a low rate of movement, *ϕ*_*i*_, back from protected to unprotected status (Figure 6).

Regardless of whether the individual is in the U or P subgroups, the flow between the SIR compartments follows the typical disease transmission process, where susceptible individuals acquire infection through contact with an infected individual, and recover after a certain time period. The force of infection, *λ*_*i*_, for susceptible individuals in political party, *i*, is given by:

,
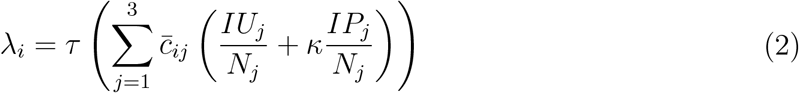

Where:

- *τ* is the baseline transmission probability per effective contact;
- 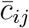is the average number of contacts per unit time that one individual in party *i* has with people in party *j*;
- 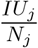 represents the proportion of party *j* in the infected-unprotected class;
- 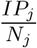 represents the proportion of party *j* in the infected-protected class;
- and *κ* is the protection scaling factor that reduces the probability of transmission for contacts that involve protected individuals.

Individuals in the infected compartment either recover (at rate *ρ*) or die (at rate *µ*). Over time, immunity wanes at rate *γ*, moving individuals from the recovered compartment back to the susceptible compartment.

To ensure comparability across groups, at the start of the simulation, we seed the infection by introducing one infected individual into each partisan group. We assume the population is fully susceptible to the pathogen at the start of the simulation, and run the simulation for one year. For all simulations, we assume the total population size is 10 million. We investigate several versions of the model to unpack the effects of partisan differences in health behaviors; the population composition of partisanship; patterns of homophily by partisanship in the contact network; and partisan responsiveness to epidemic conditions. Although we highlight differences in vaccination rates between partisan groups in Wave 6 of the survey, we do not mechanistically model the impact of vaccination, as we only had a single wave of data on vaccination at the start of the COVID-19 vaccination campaign, when the vast majority of people were still ineligible for the vaccine. These simulations are intended to provide insight into how partisan differences might affect the dynamics of a typical respiratory disease; they are not intended to exactly model COVID-19 or any other specific historical outbreak.

### Components of the SIR model that vary by party

Patterns of contact rates and mask use for each partisan group were based on group averages across all waves in the BICS data (Table 1). Because we are interested in population-level simulations rather than time or location-specific modeling, we used survey data weighted to be nationally representative and averaged across all six waves of the survey to calculate the average contact rates by party affiliation.

We chose values for the adoption of protective behavior (*π*_*i*_) and the waning protection rate (*ϕ*_*i*_) to match the general pattern of party-specific mask-wearing rates observed in the BICS data over time (Table S7).

### Homophily and Relative Size of Partisan Group

We model two additional factors that can play an important role in the structure of contacts: homophily and partisan composition. Previous research has revealed that contact networks often exhibit moderate to high levels of homophily: people who are similar to each other are more likely to have contact with one another (McPherson, Smith-Lovin and Cook, 2001). We account for the possibility of homophily by political affiliation in our model by including a parameter *β* that controls the extent to which contacts are disproportionately likely to happen within partisan groups (Currarini, Jackson and Pin, 2010). The relationship between *β* and average daily contact rates between and within partisan groups is described in detail in Supplementary Section 0.1. Briefly, when *β* = 1, contacts are independent of partisan affiliation. As *β* increases above 1, homophily increases and people interact more with others who have the same political affiliation relative to those with a different affiliation. We cannot directly measure partisan homophily from the BICS data, so we systematically vary the homophily parameter (*β* ∈ [1, 5]) and investigate how changing homophily affects the dynamic of an outbreak.

Partisan composition also affects contact structure; for example, larger Republican populations increase the likelihood of all individuals having more contacts with Republicans. We account for partisan composition in our model by including parameters that governs the share of the initial population that are Republican and Democrat. There is no typical value to use for this parameter, since partisan composition varies considerably across the United States. For example, in 2024, the two-party share of people who are registered as Republicans varied from a low of 8% in Massachusetts to a high of 80% in Wyoming (NCSL, 2025). We therefore also explore a range of values of partisan composition (Republican share 20%-70%).

## Data, Materials, and Software Availability

All study data are available at https://dataverse.harvard.edu/dataset.xhtml?persistentId=doi:10.7910/D All code for replicating the analysis are available at https://github.com/chrissoria/bics-partisan-disease-paper. A Shiny App for exploring model simulations is available at https://huggingface.co/berkeley/partisan-disease-simulator.

## Acknowledgments

C.S. received support for this research from the Greater Good Science Center Libby Fee Fellowship and the Bashir Ahmed Fellowship. A.S.M. received support for this research from the National Institute of General Medical Sciences (NIGMS) of the National Institutes of Health under award number R35GM156856. D.M.F. received support from the Hellman Foundation and the Berkeley Population Center (NICHD P2C HD 073964). A.M.D. received support from a Eunice Kennedy Shriver National Institute of Child Health and Human Development research infrastructure grant, P2C HD042828, to the Center for Studies in Demography and Ecology at the University of Washington.

## Author contributions

All authors designed the study. C.S., A.S.M., and D.M.F. analyzed data; A.M.D. provided critical feedback and reviewed the analysis. C.S. led visualization efforts and developed the companion Shiny app; all authors wrote the manuscript.

## Supporting Information

## Supporting Information Appendix

### 0.1 Key Term Definitions

#### Contacts

We define contacts as in-person physical or conversational interactions with other individuals. Conversational contacts were defined as (two-way conversation with three or more words in the physical presence of another person. This encompasses a wide range of encounters, from brief exchanges with cashiers to extended interactions with family members or fellow churchgoers.

#### Homophily

The homophily parameter *β* determines the degree of within-group interaction. When this parameter is set to 1, it implies random mixing based purely on the relative sizes of the groups. Values greater than 1 indicate a preference for in-group contacts, while values less than 1 would indicate a preference for out-group contacts (Figure S10).

#### Mask Use (Percent of Contacts with a Mask)

For respondents reporting non-zero contacts outside the household, we assessed mask-wearing behavior for up to three contacts using the question “During this contact, did you wear a face mask?” We calculate the percentage of these reported contacts during which the respondent wore a mask.

#### Protected/Unprotected Subgroups

Within each partisan group in our SIR model, individuals are further classified as either “protected” (P) or “unprotected” (U) based on their adoption of protective health behaviors such as mask-wearing. Individuals in protected subgroups have a lower probability of acquiring infection following contact with an infected individual, as well as a lower probability of transmitting infection to susceptible individuals.

#### Cumulative Incidence

The total proportion of the population that has been infected over the course of the simulation. This measure captures the overall disease burden experienced by a group or population.

#### Peak Incidence

The maximum number of new infections occurring per unit time during the outbreak. This measure summarizes the intensity of the epidemic at its most severe point and is relevant for understanding healthcare system strain.

#### Uniform Behavior Model

A simulation scenario that treats the population as homogenous, with average contact rates and mask usage values set at values observed in BICS data aggregated across all partisan groups.

#### Partisan Heterogeneity Model

A simulation scenario that incorporates partisan-specific contact rates and mask usage behaviors but assumes random mixing across partisan groups (i.e., no homophily, *β* = 1).

#### Homophilous Partisan Heterogeneity Model

A simulation scenario that incorporates both partisan-specific health behaviors and preferential within-group mixing (*β* = 5), representing a population where individuals are more likely to have contact with others of the same political affiliation.

### 0.2 Decomposing the effects of specific health behaviors and comparing partisan differences in outcomes

Although both mask use and contact patterns influence the severity of the Republican epidemic curve—resulting in higher peak prevalence, earlier peak timing, and greater cumulative mortality—contact rates play a more substantial role. Differences in mask usage alone result in 5.4% more Republican deaths relative to Democrats after one year, whereas differences in contact rates lead to a 10.4% gap (Table S8).

Incorporating both differences in mask usage and contact rates into the “Partisan Heterogeneity” model amplifies the difference in outcomes between Republicans and Democrats. Republicans exhibit approximately 16% higher death rates than Democrats after one year, growing to 17% after two years (Table S8). When accounting for the older average age of Republicans, the cumulative death disparity increases dramatically to over 75% after one year (Figure S6).

Interestingly, although Democrat behaviors are most protective in the Masks + Contacts model, peak prevalence is highest here for them (15.7%) and the time needed to reach that peak is shortest (130 days). Simultaneously, the percent Democrat deceased is smallest in this model. This occurs because, as we incorporate more Republican behaviors, which tend to be less cautious, a higher proportion of Democrat infections come from Republicans in this scenario where we do not incorporate any homophily. Given Republicans’ less cautious behavior, their group’s peak comes sooner (128 days) and thus they reach outbreak peak sooner (Table S8).

### 0.3 Temporal patterns in partisan health behaviors

Beyond these aggregate differences, temporal analysis reveals important patterns in how these behaviors evolved over the first year and half of the COVID-19 pandemic. Analysis of data across survey waves revealed that partisan contact rates and masking rates fluctuated considerably over time (Figure S1). In every survey wave, Democrats exhibited greater protective health behaviors compared to Republicans. Over time, Democrats monotonically increased their contact rates, while there was more volatility in contact rates of Republicans and Independents (Figure S3). Quantitative analysis confirmed this pattern, with Republicans showing the highest relative volatility in contact patterns (coefficient of variation = 0.26) compared to Independents (0.19) and Democrats (0.16). These differences in variance were statistically significant (Levene’s test: F = 15.47, df = 2, p *>* 0.001), indicating group-level variability across political parties (Table S12).

### 0.4 Disease Model Parameters and Equations

We develop a susceptible-infected-recovered (SIR) model that incorporates partisan differences in contact behavior and in adoption of protective behavior. The population is divided into three partisan groups, corresponding to Democrats, Republicans, and Independents; we index these groups using *i*. Within each partisan group *i*, there are four disease statuses: Susceptible, Infected, Recovered, and Deceased; and there are also two protective behavior statuses: Unprotected and Protected. All together, each partisan group has eight compartments:

- SU_*i*_, SP_*i*_: Susceptible individuals (unprotected, protected)
- IU_*i*_, IP_*i*_: Infected individuals (unprotected, protected)
- RU_*i*_, RP_*i*_: Recovered individuals (unprotected, protected)
- DU_*i*_, DP_*i*_: Deceased individuals (unprotected, protected at time of death)

At any point in time, the total population in group *i* is:

,
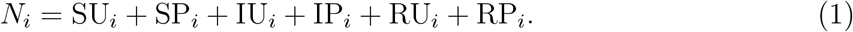

Note that deceased people are removed from the population.

#### Key Features of Model Dynamics

Model dynamics are specified as a system of ordinary differential equations; these are written out in detail in a section below and also depicted in Figure 6. Model parameters are also summarized in Table S7.

Here, we discuss two key features of our model in more detail: first, we explain how contacts work; second, we explain how contacts produce infections.

##### Contacts

In our model, contacts are a function of partisan group. The contact matrix **C** contains entries 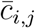, which have the average number of daily contacts that a single person in group *i* has with people in group *j*. We also call 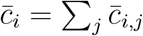 the average number of contacts that a single member of group *i* has with people in all groups.

Cross-group contacts must satisfy a balance constraint: the total number of contacts from group *i* to group *j* must equal those from group *j* to group *i*:

,
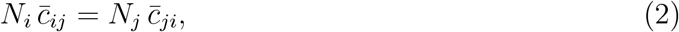

where *N*_*i*_, *N*_*j*_ are the total number of people in group *i* and *j*.

##### Homophily: Modeling preferential in-group contact

Homophily refers to a near-universal pattern found in human social networks: people are disproportionately likely to be connected to others who are similar to themselves (McPherson, Smith-Lovin and Cook, 2001; Currarini, Jackson and Pin, 2010). Previous analyses of contact networks have generally found strong evidence of homophily by age (e.g., Mossong et al. (2008)). Relatively few studies have investigated mixing in contact networks beyond age, but a few studies have found evidence of evidence of homophily by additional characteristics, including race/ethnicity and social groups such as class, sports team, work group, etc. (e.g., Pasquale et al. (2024); Glass and Glass (2008); Huang et al. (2016); Potter, Smieszek and Sailer (2015)).

We are not aware of any study that documents the extent of political homophily in inter-personal contact networks, but the widespread evidence of homophily in contact and other social networks, together with the spatial and demographic clustering of political affiliations (e.g., Brown and Enos (2021)), make it highly plausible that interpersonal contact networks are homophilous with respect to partisan identity. We therefore introduce a parameter in our disease model that controls the extent to which contacts are more likely to take place among members of the same partisan group. We adopt a parameterization of homophily inspired by Currarini, Jackson and Pin (2010).

Note that, in our model, homophily by partisan identity could arise as a result of personal preferences – but it could also result from structural factors that make it more likely for same-partisan people to encounter one another. We do not speculate about the possible origins of partisan homophily here; instead, our goal is to understand its possible consequences.

Formally, recall that *N*_*i*_ is the number of people in group *i*; let 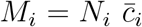 be the total number of contacts that all members of group *i* have per unit of time; and let *M* =∑_*i*_ *M*_*i*_ be the total number of contacts the entire population has per unit of time. We define the share of all contacts that is attributable to group *i* to be

,
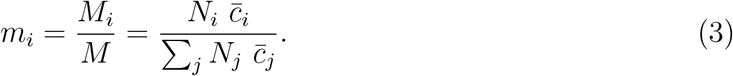

So, if we sampled a contact at random from the population and chose one of the two people involved, the chances of that person being in group *i* would be *m*_*i*_.

Let *q*_*ij*_ be the share of group *i*’s contacts that take place with people in group *j*. Then the average number of contacts someone in group *i* has with people in group *j*, 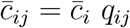.

We focus on populations with three groups, but this setup could be generalized to additional groups in other contexts. We introduce a parameter *β* ≥ 1 which controls the amount of homophily in contacts by tuning the extent to which the first group’s contacts tend to be with other members of the first group and the extent to which the second group’s contacts tend to be with other members of the second group. We cannot then independently vary contacts within the third group, as we shall see below.

Our approach takes several values as given, and then uses those values plus assumptions which we describe below to produce the matrix **C** whose entries 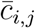 have the average number of contacts per unit time that someone in group *i* has with people in group *j*. Given:

- the population sizes of each group, *N*_1_, *N*_2_, and *N*_3_
- the average number of contacts people in each group have, 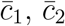, and 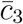
- a value of the homophily parameter *β* ≥ 1

We can calculate the matrix **C** as follows. Starting with the first group, we set

,
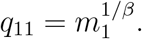

Note that when the homophily parameter *β* = 1, then *q*_11_ = *m*_1_; that is, there is no tendency for members of group 1 to have more contact with others in the same group. People in group 1 have contact with others in group 1 in proportion to the share of total contacts that group *a* represents. Thus, when *β* = 1, contacts happen at random, paying no attention to group membership. When *β >* 1, then *q*_11_ *> m*_1_ and people in group 1 are more likely to have contact with others in group 1 than would be expected at random.

We similarly use *β* to set the share of contacts that happen among members of group 2 using

,
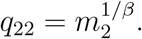

We then calculate rates of contact between all pairs of groups making simple assumptions. First, we fill in group 1’s contacts with groups 2 and 3 by assuming that they happen in proportion to each group’s share of total population contacts, *m*_2_ and *m*_3_. Specifically, we calculate

,
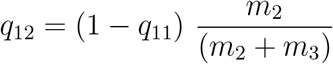

and

,
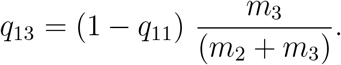

We now have a complete description of group 1’s contacts, given by the size of group 1 (*N*_1_), the average number of contacts people in group 1 have 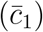, and the vector **q**_1_ = (*q*_11_, *q*_12_, *q*_13_)^*T*^ that describes the share of 1’s contacts that takes place with each group.

We will now make use of a symmetry condition that follows from the symmetry condition described above (Equation 2): for any two groups, say *i* and *j*,

,
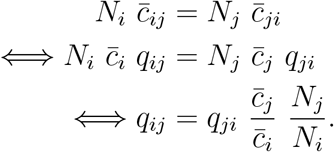

Using this symmetry condition, we can deduce the rate of contacts between group 2 and group 1:

,
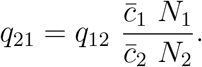

We have expressions for *q*_22_ and *q*_21_, so this means that *q*_23_ = 1 − *q*_22_ − *q*_21_, and we now know everything about contacts for group 2.

Turning to group 3, we again use the symmetry condition to deduce the rate of contacts between group 3 and group 1:

,
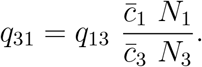

And again to deduce the rate of contacts between group 3 and group 2:

,
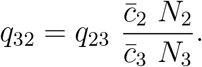

Then we deduce *q*_33_ = 1 *− q*_31_ *− q*_32_.

Finally, we can calculate the entries of the contact matrix **C**, which has the average number of daily contacts between groups, using 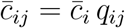

#### Force of Infection

The force of infection for group *i, λ*_*i*_, governs the rate at which susceptibles in group *i* become infected. *λ*_*i*_ depends on average daily contact rates that individuals in group *i* have with individuals in all groups, including *i*, and the infection prevalence for each group:

,
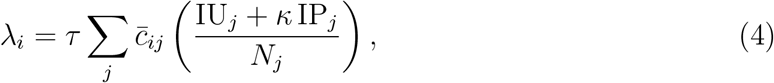

where *κ* is a parameter capturing the efficacy of protective behavior and 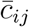 is, as defined in the previous section, the average number of contacts that a single member of group *i* has with people in group *j*.

In Equation 4, we assume that (i) the 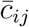 contacts happen with unprotected and protected members of group *j* in proportion to the population composition of group *j*; and (ii) that protected and unprotected members of group *j* have the same average number of contacts, but that contacts with protected members are less effective at transmitting the infection.

#### SIR Model Equations

Here, we write out the full set of ordinary differential equations that describe the rate of movement in an out of the compartments in our SIR model.

##### Susceptible Unprotected (SU_*i*_)

,
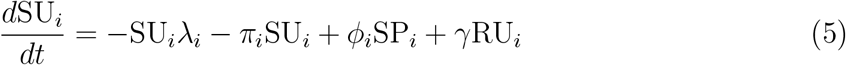

##### Susceptible Protected (SP_*i*_)

,
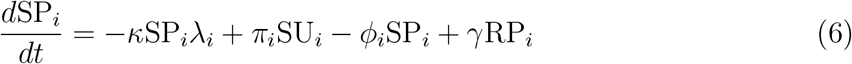

##### Infected Unprotected (IU_*i*_)

,
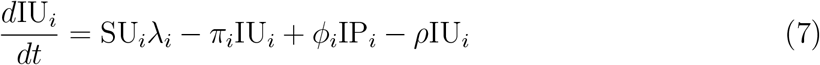

##### Infected Protected (IP_*i*_)

,
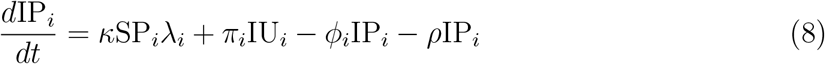

##### Recovered Unprotected (RU_*i*_)

,
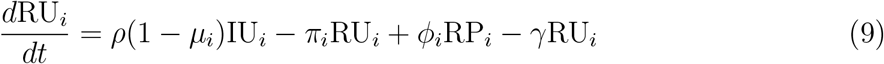

##### Recovered Protected (RP_*i*_)

,
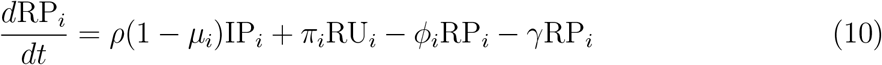

##### Deceased Unprotected (DU_*i*_)

,
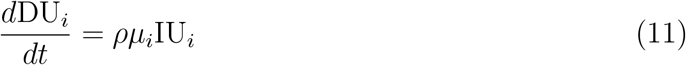

##### Deceased Protected (DP_*i*_)

,
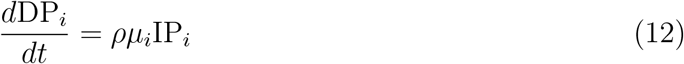

### 0.5 Age-adjusted infection fatality rates

In our main simulations, we assume a uniform infection fatality rate (*µ*) across all partisan groups. However, Republicans in the BICS sample are older on average than Democrats and Independents (Table S3), and many respiratory infectious diseases exhibit strong age-mortality gradients. To examine how age composition differences might affect partisan mortality disparities, we conducted a sensitivity analysis using age-specific infection fatality rates.

We calculated group-specific infection fatality rates using the meta-regression formula from Levin et al. (2020), which estimated age-specific COVID-19 infection fatality rates:

,
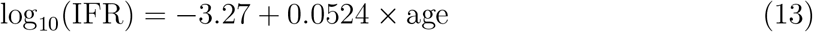

Using the mean age of each partisan group in the BICS data, we derived the following group-specific infection fatality rates: *µ*_*R*_ = 0.0150 for Republicans, *µ*_*I*_ = 0.0119 for Independents, and *µ*_*D*_ = 0.0099 for Democrats. These rates reflect the older average age of Republicans relative to Democrats in our sample.

In the SIR model, the infection fatality rate *µ*_*i*_ determines the proportion of infected individuals in group *i* who die rather than recover. Specifically, infected individuals leave the infected compartment at rate *ρ*, with a fraction *µ*_*i*_ dying and a fraction (1*−µ*_*i*_) recovering:

,
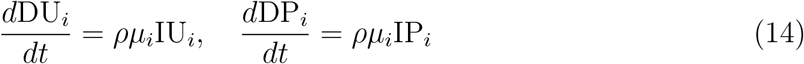

When we incorporate these age-adjusted fatality rates into our simulations (Figure S6), the mortality disparity between Republicans and Democrats increases dramatically—from approximately 16% higher Republican mortality (with uniform *µ*) to over 75% higher mortality. This amplification occurs because Republicans face a “double burden”: their higher contact rates and lower adoption of protective behaviors lead to higher infection rates, and their older age distribution means a larger fraction of those infections prove fatal.

### Supporting Tables

**Table S1:** Vaccination Behavior by Political Affiliation (Wave 6). Self-reported COVID-19 vaccination status and intentions from BICS Wave 6 (May 2021), weighted for national representativeness. Among unvaccinated respondents, the table shows the percentage who reported they would or would not get the vaccine when available. Republicans showed lower vaccination rates (57%) compared to Democrats (74%), and among unvaccinated respondents, a substantially higher percentage of Republicans (73%) reported they would not get the vaccine compared to Democrats (48%).

|  | % Vaccinated | Not Vaccinated |  |  |
|---|---|---|---|---|
| Party |  | % Will Get Vaccine | % Won't Get Vaccine | N |
| Republican | 57 | 26 | 73 | 1411 |
| Independent | 61 | 31 | 69 | 1295 |
| Democrat | 74 | 52 | 48 | 2288 |

**Table S2:** Reported Pandemic Behaviors by Political Affiliation Across Waves. Unadjusted weighted averages of pandemic health behaviors by political affiliation across six BICS survey waves (April 2020 – May 2021). Total contacts and non-household contacts are reported as previous-day averages; mask use is the percentage of contacts during which respondents wore a mask; vaccination rates are from Wave 6 only (May 2021). These estimates are weighted for national representativeness but are not adjusted for demographic or geographic covariates. Republicans consistently reported more daily contacts and lower mask usage than Democrats across all waves, with the largest contact gap occurring in September 2020 (8.64 vs. 5.19 total contacts). Contact rates for all groups generally increased over time, while mask usage peaked in late 2020 before declining in 2021.

| Party | Behaviors | Apr, ’20 | Jun, ’20 | Sept, ’20’ | Dec, ’20’ | Feb, ’21’ | May, ’21’ |
|---|---|---|---|---|---|---|---|
| Democrat | Total Contacts | 3.50 | 4.92 | 5.19 | 5.23 | 5.64 | 6.14 |
|  | Non-Household Contacts | 1.79 | 3.10 | 3.47 | 3.37 | 3.72 | 4.28 |
|  | % Contacts with a Mask | 59.60 | 59.71 | 69.70 | 72.71 | 72.30 | 59.85 |
|  | % Vaccinated |  |  |  |  |  | 74.00 |
| Independent | Total Contacts | 4.12 | 4.93 | 6.04 | 5.47 | 4.84 | 7.23 |
|  | Non-Household Contacts | 2.26 | 2.95 | 4.14 | 3.77 | 3.17 | 5.35 |
|  | % Contacts with a Mask | 50.54 | 44.48 | 66.56 | 61.47 | 64.46 | 46.67 |
|  | % Vaccinated |  |  |  |  |  | 61.00 |
| Republican | Total Contacts | 4.40 | 5.67 | 8.64 | 5.78 | 5.95 | 7.71 |
|  | Non-Household Contacts | 2.36 | 3.82 | 6.62 | 3.89 | 4.09 | 5.90 |
|  | % Contacts with a Mask | 47.33 | 51.81 | 64.70 | 61.42 | 57.63 | 45.14 |
|  | % Vaccinated |  |  |  |  |  | 57.00 |
| Total | Total Contacts | 3.92 | 5.17 | 6.59 | 5.45 | 5.54 | 6.91 |
|  | Non-Household Contacts | 2.07 | 3.30 | 4.72 | 3.62 | 3.70 | 5.06 |
|  | % Contacts with a Mask | 53.38 | 53.03 | 67.05 | 66.56 | 66.01 | 51.85 |
|  | % Vaccinated |  |  |  |  |  | 65.00 |

**Table S3:** Demographic Characteristics by Political Affiliation. Unadjusted weighted averages of demographic characteristics by political affiliation from pooled BICS survey data (April 2020 – May 2021). All measures are self-reported; college degree refers to a four-year degree. Republicans in the sample were older on average (50.2 years vs. 46.1 for Democrats), more likely to be male (51.2% vs. 46.6%), more likely to be White (89.8% vs. 69.9%), and less likely to live in metropolitan areas (82.0% vs. 87.4%). These compositional differences are accounted for in the regression analyses presented in the main text.

| Variable | Democrats | Independents | Republicans |
|---|---|---|---|
| Average Age | 46.1 | 46.4 | 50.2 |
| Average Household Size | 2.9 | 2.9 | 2.9 |
| % Male | 46.6 | 49.7 | 51.2 |
| % White | 69.9 | 78.1 | 89.8 |
| % Hispanic | 20.2 | 13.7 | 11.8 |
| % Living in Metro | 87.4 | 85.4 | 82.0 |
| % College Degree | 37.2 | 29.2 | 31.4 |
| % Employed | 59.8 | 54.5 | 58.1 |

**Table S4:** Association between Political Party and the Number of Reported Contacts. Linear regression models predicting total daily contacts from BICS survey data (April 2020 – May 2021). Model 1 includes party affiliation and survey wave controls. Model 2 adds demographic covariates (age, race, ethnicity, gender, education, employment, household size). Model 3 adds geographic and policy controls (metropolitan area, congressional district percent Democrat, COVID-19 incidence rate, mask mandate status, and state fixed effects). Republicans reported significantly more contacts than Democrats across all model specifications, with an adjusted difference of approximately 1.1 additional daily contacts in the fully controlled model (Model 3). Standard errors in parentheses.

|  | *Dependent variable:* |  |  |
|---|---|---|---|
|  | Total Contacts |  |  |
|  | Model 1 | Model 2 | Model 3 |
|  | (1) | (2) | (3) |
| Independent Party (ref: Democrat) | 0.432<sup>**</sup> | 0.452<sup>**</sup> | 0.362<sup>*</sup> |
|  | (0.192) | (0.194) | (0.202) |
| Republican Party (ref: Democrat) | 1.364<sup>***</sup> | 1.247<sup>***</sup> | 1.125<sup>***</sup> |
|  | (0.178) | (0.184) | (0.192) |
| Prefer Not to Answer (ref: Democrat) | 1.522<sup>***</sup> | 1.549<sup>***</sup> | 1.800<sup>***</sup> |
|  | (0.309) | (0.328) | (0.338) |
| Wave 2 (ref: Wave 1) | 1.098<sup>***</sup> | 1.074<sup>***</sup> | 0.890<sup>**</sup> |
|  | (0.313) | (0.360) | (0.372) |
| Wave 3 (ref: Wave 1) | 2.427<sup>***</sup> | 2.408<sup>***</sup> | 2.150<sup>***</sup> |
|  | (0.292) | (0.344) | (0.359) |
| Wave 4 (ref: Wave 1) | 1.586<sup>***</sup> | 1.677<sup>***</sup> | 1.532<sup>***</sup> |
|  | (0.295) | (0.345) | (0.356) |
| Wave 5 (ref: Wave 1) | 1.723<sup>***</sup> | 1.692<sup>***</sup> | 1.724<sup>***</sup> |
|  | (0.296) | (0.347) | (0.365) |
| Wave 6 (ref: Wave 1) | 3.047<sup>***</sup> | 2.925<sup>***</sup> | 2.843<sup>***</sup> |
|  | (0.269) | (0.325) | (0.338) |
| Weekend Interview | −0.227 | −0.157 | −0.169 |
|  | (0.149) | (0.150) | (0.155) |
| Age Group: [30,45) (ref: [18,30) |  | 0.015 | −0.181 |
|  |  | (0.221) | (0.229) |
| Age Group: [45,55) (ref: [18,30) |  | 0.471<sup>*</sup> | 0.386 |
|  |  | (0.251) | (0.261) |
| Age Group: [55,65) (ref: [18,30) |  | −0.584<sup>**</sup> | −0.589<sup>**</sup> |
|  |  | (0.263) | (0.271) |
| Age Group: [65,100] (ref: [18,30) |  | −1.246<sup>***</sup> | −1.256<sup>***</sup> |
|  |  | (0.259) | (0.271) |
| Race: Black (ref: White) |  | −0.754<sup>***</sup> | −0.706<sup>***</sup> |
|  |  | (0.242) | (0.255) |
| Race: Asian (ref: White) |  | −1.823<sup>***</sup> | −1.700<sup>***</sup> |
|  |  | (0.354) | (0.363) |
| Race: Other/Mixed (ref: White) |  | −0.711<sup>**</sup> | −0.598<sup>*</sup> |
|  |  | (0.338) | (0.349) |
| Hispanic |  | 0.116 | −0.014 |
|  |  | (0.160) | (0.169) |
| Male |  | 0.191 | 0.236 |
|  |  | (0.152) | (0.158) |
| High School Graduate (ref: College Grad) |  | 0.217 | 0.128 |
|  |  | (0.171) | (0.178) |
| Less than High School (ref: College Grad) |  | −0.014 | −0.779<sup>*</sup> |
|  |  | (0.424) | (0.447) |
| Employed |  | 2.580<sup>***</sup> | 2.612<sup>***</sup> |
|  |  | (0.171) | (0.178) |
| Household Size |  | 0.819<sup>***</sup> | 0.794<sup>***</sup> |
|  |  | (0.047) | (0.048) |
| Metro Area |  |  | −0.558<sup>**</sup> |
|  |  |  | (0.268) |
| CD Percent Democrat |  |  | −0.081 |
|  |  |  | (0.532) |
| Log Previous Week Incidence Rate |  |  | −0.147 |
|  |  |  | (0.126) |
| Less Strict Mask Mandate (ref: Strict Mandate) |  |  | 0.939<sup>***</sup> |
|  |  |  | (0.338) |
| Unspecified Mask Mandate (ref: Strict Mandate) |  |  | 0.496 |
|  |  |  | (0.539) |
| No Mask Mandate (ref: Strict Mandate) |  |  | −0.127 |
|  |  |  | (0.401) |
| Constant | 3.574<sup>***</sup> | 0.050 | 0.416 |
|  | (0.274) | (0.454) | (0.708) |
| Observations | 18,637 | 17,946 | 16,677 |
| R<sup>2</sup> | 0.013 | 0.062 | 0.072 |
| Adjusted R<sup>2</sup> | 0.012 | 0.061 | 0.067 |
| Residual Std. Error | 9.982 (df = 18621) | 9.859 (df = 17917) | 9.776 (df = 16593) |
| F Statistic | 16.695<sup>***</sup> (df = 15; 18621) | 42.531<sup>***</sup> (df = 28; 17917) | 15.412<sup>***</sup> (df = 83; 16593) |

*Note:* <sup>\*</sup>p<0.1; <sup>\*\*</sup>p<0.05; <sup>\*\*\*</sup>p<0.01

**Table S5:**
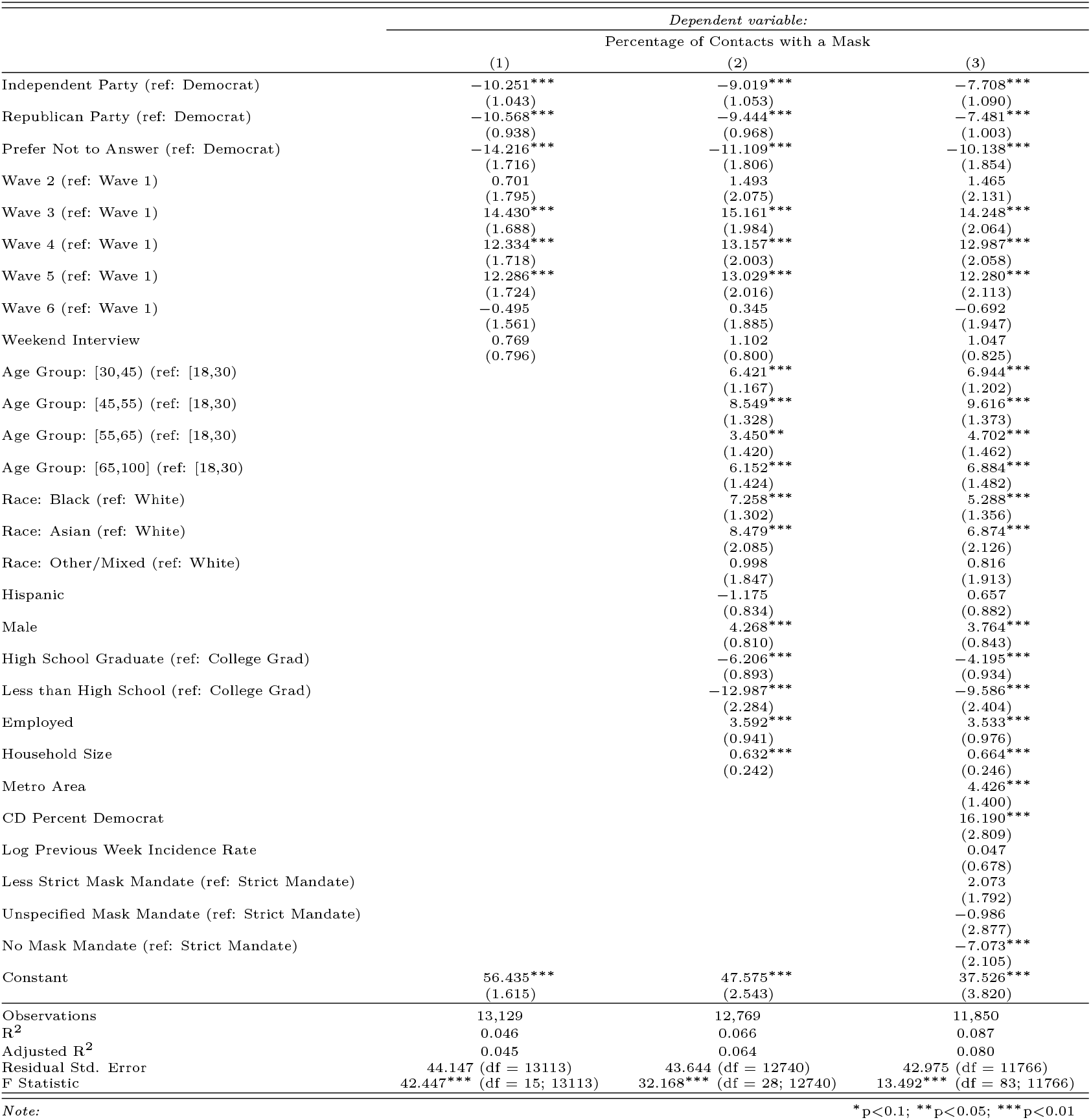
Association between Political Party and Reported Percentage of Contacts with a Mask. Linear regression models predicting the percentage of contacts during which respondents wore a mask, from BICS survey data (April 2020 – May 2021). Model 1 includes party affiliation and survey wave controls. Model 2 adds demographic covariates (age, race, ethnicity, gender, education, employment, household size). Model 3 adds geographic and policy controls (metropolitan area, congressional district percent Democrat, COVID-19 incidence rate, mask mandate status, and state fixed effects). Republicans reported significantly lower mask usage than Democrats across all model specifications, with an adjusted difference of approximately 7.5 percentage points lower mask use in the fully controlled model (Model 3). Standard errors in parentheses.

**Table S6:**
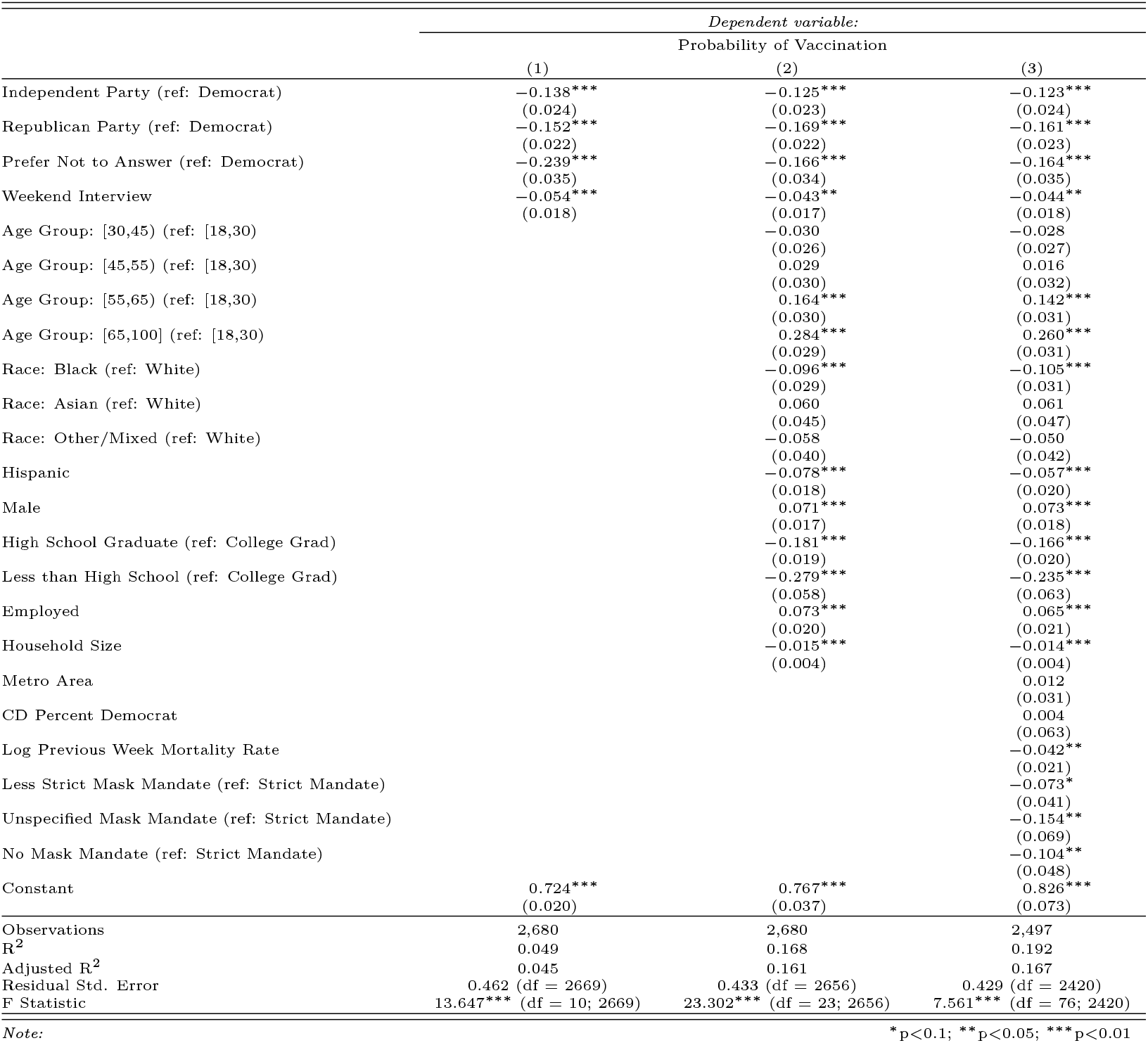
Association between Political Party and Reported Vaccination. Linear regression models predicting self-reported COVID-19 vaccination status from BICS Wave 6 data (May 2021). Vaccination data were only collected in Wave 6, when vaccines had become accessible to the general public. Model 1 includes party affiliation and survey day controls. Model 2 adds demographic covariates (age, race, ethnicity, gender, education, employment, household size). Model 3 adds geographic and policy controls (metropolitan area, congressional district percent Democrat, COVID-19 mortality rate, mask mandate status, and state fixed effects). Republicans were significantly less likely to be vaccinated than Democrats across all model specifications, with an adjusted difference of approximately 16 percentage points lower vaccination probability in the fully controlled model (Model 3). Standard errors in parentheses.

|  | (1) | (2) | (3) |
|---|---|---|---|
|  | *Dependent variable:* |  |  |
|  | Probability of Vaccination |  |  |
|  | (1) | (2) | (3) |
|---|---|---|---|
| Independent Party (ref: Democrat) | −0.138<sup>***</sup> | −0.125<sup>***</sup> | −0.123<sup>***</sup> |
|  | (0.024) | (0.023) | (0.024) |
| Republican Party (ref: Democrat) | −0.152<sup>***</sup> | −0.169<sup>***</sup> | −0.161<sup>***</sup> |
|  | (0.022) | (0.022) | (0.023) |
| Prefer Not to Answer (ref: Democrat) | −0.239<sup>***</sup> | −0.166<sup>***</sup> | −0.164<sup>***</sup> |
|  | (0.035) | (0.034) | (0.035) |
| Weekend Interview | −0.054<sup>***</sup> | −0.043<sup>**</sup> | −0.044<sup>**</sup> |
|  | (0.018) | (0.017) | (0.018) |
| Age Group: [30,45) (ref: [18,30) |  | −0.030 | −0.028 |
|  |  | (0.026) | (0.027) |
| Age Group: [45,55) (ref: [18,30) |  | 0.029 | 0.016 |
|  |  | (0.030) | (0.032) |
| Age Group: [55,65) (ref: [18,30) |  | 0.164<sup>***</sup> | 0.142<sup>***</sup> |
|  |  | (0.030) | (0.031) |
| Age Group: [65,100] (ref: [18,30) |  | 0.284<sup>***</sup> | 0.260<sup>***</sup> |
|  |  | (0.029) | (0.031) |
| Race: Black (ref: White) |  | −0.096<sup>***</sup> | −0.105<sup>***</sup> |
|  |  | (0.029) | (0.031) |
| Race: Asian (ref: White) |  | 0.060 | 0.061 |
|  |  | (0.045) | (0.047) |
| Race: Other/Mixed (ref: White) |  | −0.058 | −0.050 |
|  |  | (0.040) | (0.042) |
| Hispanic |  | −0.078<sup>***</sup> | −0.057<sup>***</sup> |
|  |  | (0.018) | (0.020) |
| Male |  | 0.071<sup>***</sup> | 0.073<sup>***</sup> |
|  |  | (0.017) | (0.018) |
| High School Graduate (ref: College Grad) |  | −0.181<sup>***</sup> | −0.166<sup>***</sup> |
|  |  | (0.019) | (0.020) |
| Less than High School (ref: College Grad) |  | −0.279<sup>***</sup> | −0.235<sup>***</sup> |
|  |  | (0.058) | (0.063) |
| Employed |  | 0.073<sup>***</sup> | 0.065<sup>***</sup> |
|  |  | (0.020) | (0.021) |
| Household Size |  | −0.015<sup>***</sup> | −0.014<sup>***</sup> |
|  |  | (0.004) | (0.004) |
| Metro Area |  |  | 0.012 |
|  |  |  | (0.031) |
| CD Percent Democrat |  |  | 0.004 |
|  |  |  | (0.063) |
| Log Previous Week Mortality Rate |  |  | −0.042<sup>**</sup> |
|  |  |  | (0.021) |
| Less Strict Mask Mandate (ref: Strict Mandate) |  |  | −0.073<sup>*</sup> |
|  |  |  | (0.041) |
| Unspecified Mask Mandate (ref: Strict Mandate) |  |  | −0.154<sup>**</sup> |
|  |  |  | (0.069) |
| No Mask Mandate (ref: Strict Mandate) |  |  | −0.104<sup>**</sup> |
|  |  |  | (0.048) |
| Constant | 0.724<sup>***</sup> | 0.767<sup>***</sup> | 0.826<sup>***</sup> |
|  | (0.020) | (0.037) | (0.073) |
| Observations | 2,680 | 2,680 | 2,497 |
| R<sup>2</sup> | 0.049 | 0.168 | 0.192 |
| Adjusted R<sup>2</sup> | 0.045 | 0.161 | 0.167 |
| Residual Std. Error | 0.462 (df = 2669) | 0.433 (df = 2656) | 0.429 (df = 2420) |
| F Statistic | 13.647<sup>***</sup> (df = 10; 2669) | 23.302<sup>***</sup> (df = 23; 2656) | 7.561<sup>***</sup> (df = 76; 2420) |

*Note:* <sup>\*</sup>p<0.1; <sup>\*\*</sup>p<0.05; <sup>\*\*\*</sup>p<0.01

**Table S7:**
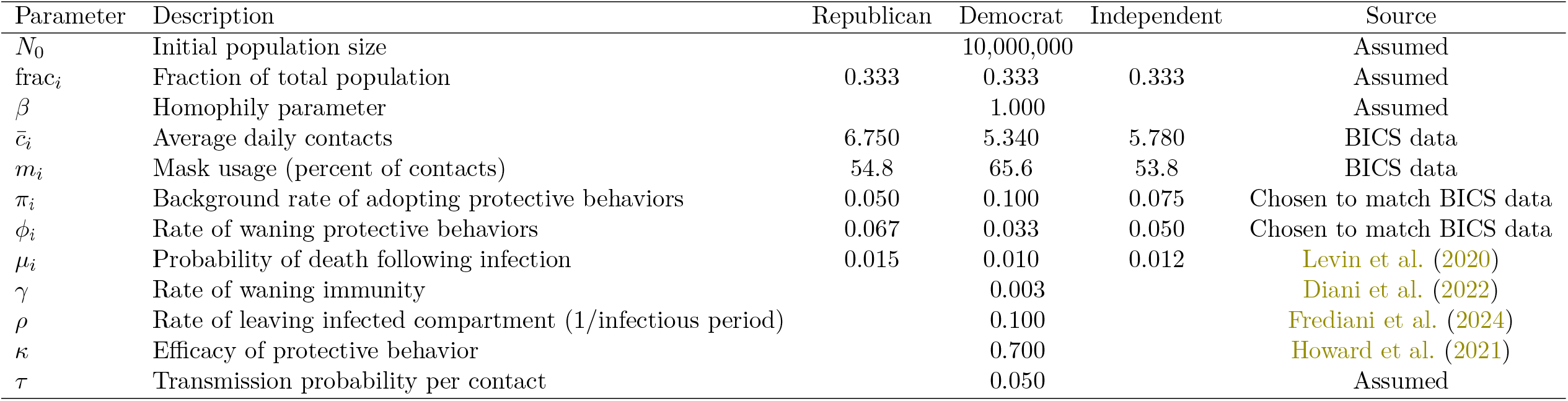
Partisan Model Parameters. Parameter values used in the three-party SIR epidemiological model. Partisan-specific contact rates 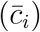 and mask usage (*m*_*i*_) are derived from pooled BICS survey data (April 2020 – May 2021). Rates of adopting (*π*_*i*_) and waning (*ϕ*_*i*_) protective behaviors were chosen to match temporal patterns in mask usage observed in the BICS data. Infection fatality rates (*µ*_*i*_) are age-adjusted using estimates from Levin et al. (2020), which found that infection fatality rates increase exponentially with age. Epidemiological parameters (waning immunity *γ*, infectious period *ρ*, mask efficacy *κ*) are drawn from published literature. Baseline simulations assume equal population shares across partisan groups and random mixing (*β* = 1). Because partisan mixing patterns could not be directly measured from BICS data and reliable estimates were not available in the literature, we vary the homophily parameter (*β*) across a range of values to explore how within-group contact preferences affect disease outcomes.

| Parameter | Description | Republican | Democrat | Independent | Source |
|---|---|---|---|---|---|
| N<sub>0</sub> | Initial population size | 10,000,000 |  |  | Assumed |
| frac<sub>i</sub> | Fraction of total population | 0.333 | 0.333 | 0.333 | Assumed |
| β | Homophily parameter | 1.000 |  |  | Assumed |
| c̄<sub>i</sub> | Average daily contacts | 6.750 | 5.340 | 5.780 | BICS data |
| m<sub>i</sub> | Mask usage (percent of contacts) | 54.8 | 65.6 | 53.8 | BICS data |
| π<sub>i</sub> | Background rate of adopting protective behaviors | 0.050 | 0.100 | 0.075 | Chosen to match BICS data |
| ϕ<sub>i</sub> | Rate of waning protective behaviors | 0.067 | 0.033 | 0.050 | Chosen to match BICS data |
| μ<sub>i</sub> | Probability of death following infection | 0.015 | 0.010 | 0.012 | Levin et al. (2020) |
| γ | Rate of waning immunity | 0.003 |  |  | Diani et al. (2022) |
| ρ | Rate of leaving infected compartment (1/infectious period) | 0.100 |  |  | Frediani et al. (2024) |
| κ | Efficacy of protective behavior | 0.700 |  |  | Howard et al. (2021) |
| τ | Transmission probability per contact | 0.050 |  |  | Assumed |

**Table S8:** Comparison of Key Outcomes Across Partisan Behavior Models. Results from three-party SIR simulations comparing disease outcomes under different behavioral assumptions. “Uniform Behavior” assumes population-average contact rates and mask usage with no partisan differences. “Lowest Transmission Behavior” assumes all groups adopt the most protective behaviors observed in the data (lowest contact rates, highest mask usage). “Partisan Heterogeneity” incorporates observed partisan differences in mask usage only, contact rates only, or both (Mask + Contacts). All simulations assume random mixing (*β* = 1) and an evenly split population (33% Republican, 33% Democrat, 33% Independent). Full outbreak size and cumulative deceased are shown at 1-year and 2-year time points. Peak magnitude and days to reach peak reflect the maximum infection rate during the simulation. Rates are presented per 10,000 people.

|  | Full Outbreak Size |  | Peak |  | Cumulative Deceased |  |
|---|---|---|---|---|---|---|
| Behavior Differences | 1 Year | 2 Years | Magnitude | Days to Reach | 1 Year | 2 Years |
| Uniform Behavior |  |  |  |  |  |  |
| Total | 8,994 | 9,181 | 1,635 | 137 | 110 | 113 |
| Republican | 2,998 | 9,181 | 1,635 | 137 | 37 | 113 |
| Independent | 2,998 | 9,181 | 1,635 | 137 | 37 | 113 |
| Democrat | 2,998 | 9,181 | 1,635 | 137 | 37 | 113 |
| Lowest Transmission Behavior |  |  |  |  |  |  |
| Total | 7,463 | 7,536 | 904 | 215 | 91 | 93 |
| Republican | 2,488 | 7,536 | 904 | 215 | 30 | 93 |
| Independent | 2,488 | 7,536 | 904 | 215 | 30 | 93 |
| Democrat | 2,488 | 7,536 | 904 | 215 | 30 | 93 |
| Partisan Heterogeneity |  |  |  |  |  |  |
| Mask Usage |  |  |  |  |  |  |
| Total | 9,013 | 9,213 | 1,653 | 136 | 111 | 113 |
| Republican | 3,077 | 9,440 | 1,706 | 135 | 46 | 142 |
| Independent | 3,003 | 9,208 | 1,651 | 136 | 36 | 109 |
| Democrat | 2,933 | 8,991 | 1,603 | 136 | 29 | 89 |
| Contacts |  |  |  |  |  |  |
| Total | 8,972 | 9,183 | 1,648 | 135 | 111 | 114 |
| Republican | 3,150 | 9,683 | 1,763 | 134 | 47 | 145 |
| Independent | 2,962 | 9,092 | 1,627 | 135 | 35 | 108 |
| Democrat | 2,860 | 8,776 | 1,558 | 135 | 28 | 87 |
| Mask + Contacts |  |  |  |  |  |  |
| Total | 9,023 | 9,326 | 1,710 | 129 | 110 | 114 |
| Republican | 3,237 | 10,067 | 1,881 | 128 | 40 | 124 |
| Independent | 2,980 | 9,252 | 1,689 | 130 | 36 | 114 |
| Democrat | 2,806 | 8,661 | 1,569 | 130 | 34 | 106 |

**Table S9:** Comparison of Key Outcomes Across Partisan Behavior Models in a Republican Majority Population. Results from three-party SIR simulations with a Republican majority population composition (70% Republican, 20% Democrat, 10% Independent). “Uniform Behavior” assumes population-average contact rates and mask usage with no partisan differences and random mixing (*β* = 1). “Partisan Differences” incorporates observed partisan differences in contact rates and mask usage with random mixing (*β* = 1). “Moderate Homophily” and “High Homophily” add increasing levels of within-group contact preference (*β* = 3 and *β* = 5, respectively). Full outbreak size (FOS) and cumulative deceased are shown at the 1-year time point. Peak magnitude and days to reach peak reflect the maximum infection rate during the simulation. Rates are presented per 10,000 people.

|  | Full Outbreak Size | Peak |  | Cumulative Deceased |
|---|---|---|---|---|
| Behavior Differences | FOS | Magnitude | Days to Reach | Deceased: 365 |
| Uniform Behavior |  |  |  |  |
| Total | 9,360 | 1,869 | 121 | 114 |
| Republican | 3,120 | 1,869 | 121 | 38 |
| Independent | 3,120 | 1,869 | 121 | 38 |
| Democrat | 3,120 | 1,869 | 121 | 38 |
| Partisan Differences |  |  |  |  |
| Total | 9,778 | 2,146 | 106 | 119 |
| Republican | 7,071 | 2,244 | 106 | 86 |
| Independent | 936 | 2,025 | 107 | 11 |
| Democrat | 1,771 | 1,887 | 108 | 22 |
| Partisan Differences (Moderate Homophily) |  |  |  |  |
| Total | 9,806 | 2,133 | 103 | 119 |
| Republican | 7,239 | 2,340 | 102 | 88 |
| Independent | 899 | 1,866 | 107 | 11 |
| Democrat | 1,668 | 1,693 | 108 | 20 |
| Partisan Differences (High Homophily) |  |  |  |  |
| Total | 9,833 | 2,098 | 102 | 119 |
| Republican | 7,320 | 2,382 | 100 | 89 |
| Independent | 892 | 1,790 | 108 | 11 |
| Democrat | 1,622 | 1,567 | 110 | 20 |

**Table S10:** Comparison of Key Outcomes Across Homophily Scenarios. Results from three-party SIR simulations with an evenly split population composition (33% Republican, 33% Democrat, 33% Independent) and observed partisan differences in contact rates and mask usage. “Random Mixing” assumes no preferential within-group contact (*β* = 1). “Moderate Homophily” and “High Homophily” add increasing levels of within-group contact preference (*β* = 3 and *β* = 5, respectively). Full outbreak size and cumulative deceased are shown at 1-year and 2-year time points. Peak magnitude and days to reach peak reflect the maximum infection rate during the simulation. Rates are presented per 10,000 people.

|  | Full Outbreak Size |  | Peak |  | Cumulative Deceased |  |
|---|---|---|---|---|---|---|
| Behavior Differences | 1 Year | 2 Years | Magnitude | Days to Reach | 1 Year | 2 Years |
| Random Mixing |  |  |  |  |  |  |
| Total | 9,023 | 9,326 | 1,710 | 129 | 110 | 114 |
| Republican | 9,710 | 10,067 | 1,881 | 128 | 119 | 124 |
| Independent | 8,940 | 9,252 | 1,689 | 130 | 109 | 114 |
| Democrat | 8,419 | 8,661 | 1,569 | 130 | 103 | 106 |
| Moderate Homophily |  |  |  |  |  |  |
| Total | 8,984 | 9,477 | 1,708 | 121 | 110 | 116 |
| Republican | 10,025 | 10,711 | 2,035 | 118 | 122 | 131 |
| Independent | 8,882 | 9,364 | 1,695 | 123 | 109 | 115 |
| Democrat | 8,046 | 8,356 | 1,478 | 125 | 98 | 103 |
| High Homophily |  |  |  |  |  |  |
| Total | 8,986 | 9,636 | 1,667 | 117 | 110 | 118 |
| Republican | 10,193 | 11,197 | 2,123 | 111 | 124 | 137 |
| Independent | 8,883 | 9,484 | 1,689 | 119 | 109 | 116 |
| Democrat | 7,883 | 8,226 | 1,402 | 122 | 96 | 101 |

**Table S11:**
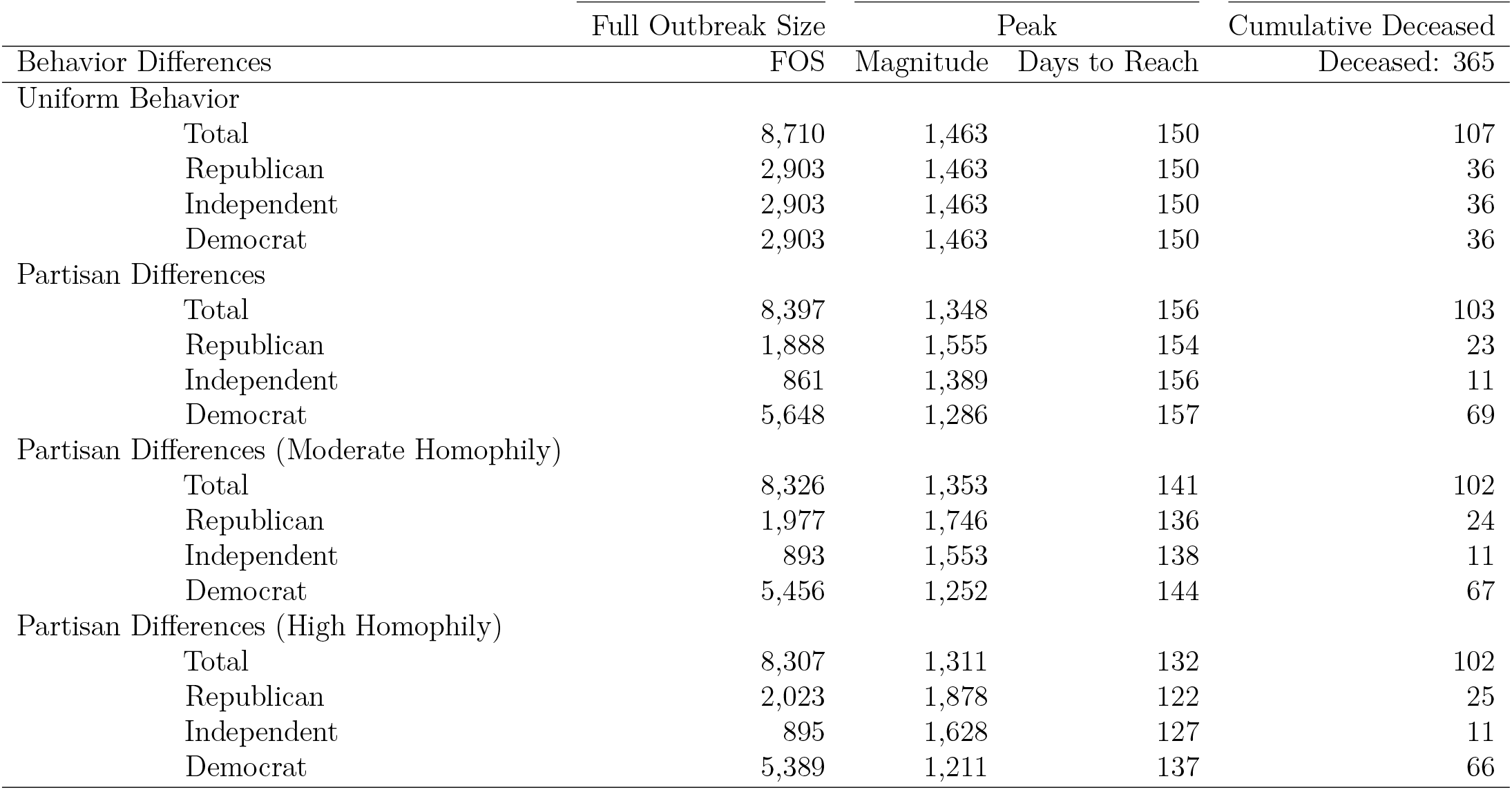
Comparison of Key Outcomes Across Partisan Behavior Models in a Democrat Majority Population. Results from three-party SIR simulations with a Democrat majority population composition (20% Republican, 70% Democrat, 10% Independent). “Uniform Behavior” assumes population-average contact rates and mask usage with no partisan differences and random mixing (*β* = 1). “Partisan Differences” incorporates observed partisan differences in contact rates and mask usage with random mixing (*β* = 1). “Moderate Homophily” and “High Homophily” add increasing levels of within-group contact preference (*β* = 3 and *β* = 5, respectively). Full outbreak size (FOS) and cumulative deceased are shown at the 1-year time point. Peak magnitude and days to reach peak reflect the maximum infection rate during the simulation. Rates are presented per 10,000 people.

**Table S12:** Volatility Measures and Levene’s Test for Total Contacts by Political Party. Measures of variability in average daily contact rates across six BICS survey waves (April 2020 – May 2021). Republicans showed the highest volatility in contact patterns (coefficient of variation = 0.26) compared to Independents (0.19) and Democrats (0.16). Levene’s test confirms that these differences in variance are statistically significant (F = 15.47, df = 2, p *<* 0.001), indicating that Republicans’ contact behaviors fluctuated more over time than those of other partisan groups. *Significant at p *<* 0.001.

| Political Party | Standard Deviation | Variance | Coefficient of Variation | Mean Contacts |
|---|---|---|---|---|
| Republican | 1.62 | 2.62 | 0.26 | 6.31 |
| Democrat | 0.81 | 0.66 | 0.16 | 5.14 |
| Independent | 1.04 | 1.07 | 0.19 | 5.48 |
| Levene Test | F = 15.471 | df = 2 |  |  |
| p-value |  | 0 | Significant* |  |

### Supporting Figures

**Figure S1:**
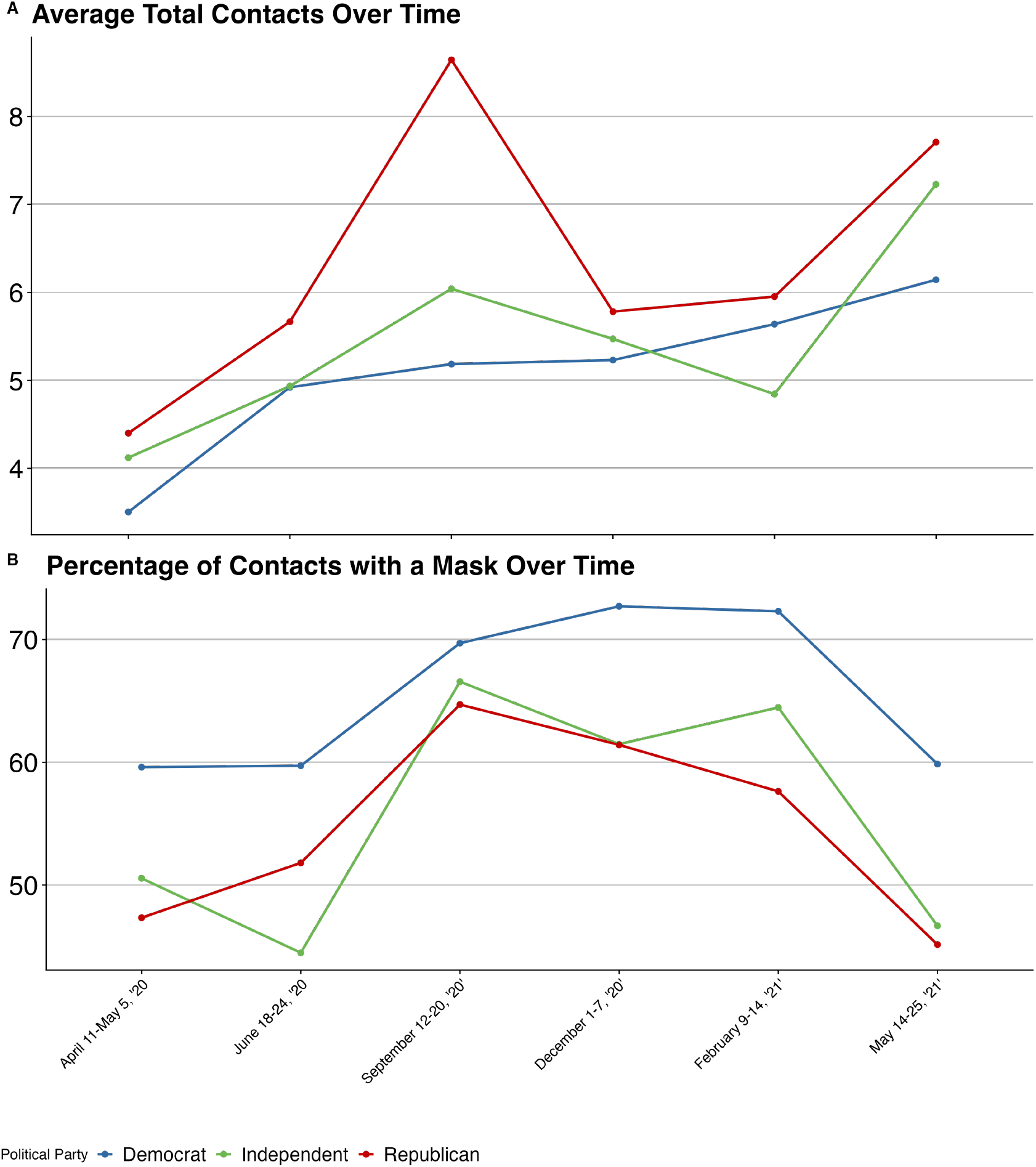
Partisan Health Behaviors Over Time. Panel (A) shows average daily in-person contacts by partisan affiliation across six BICS survey waves (April 2020 – May 2021). Panel (B) shows the percentage of contacts involving mask use by partisan affiliation over the same period. Republicans (red), Democrats (blue), and Independents (green) are shown with 95% confidence intervals. Contact rates and masking behaviors fluctuated over time in response to pandemic conditions, with persistent partisan differences throughout the study period.

**Figure S2:**
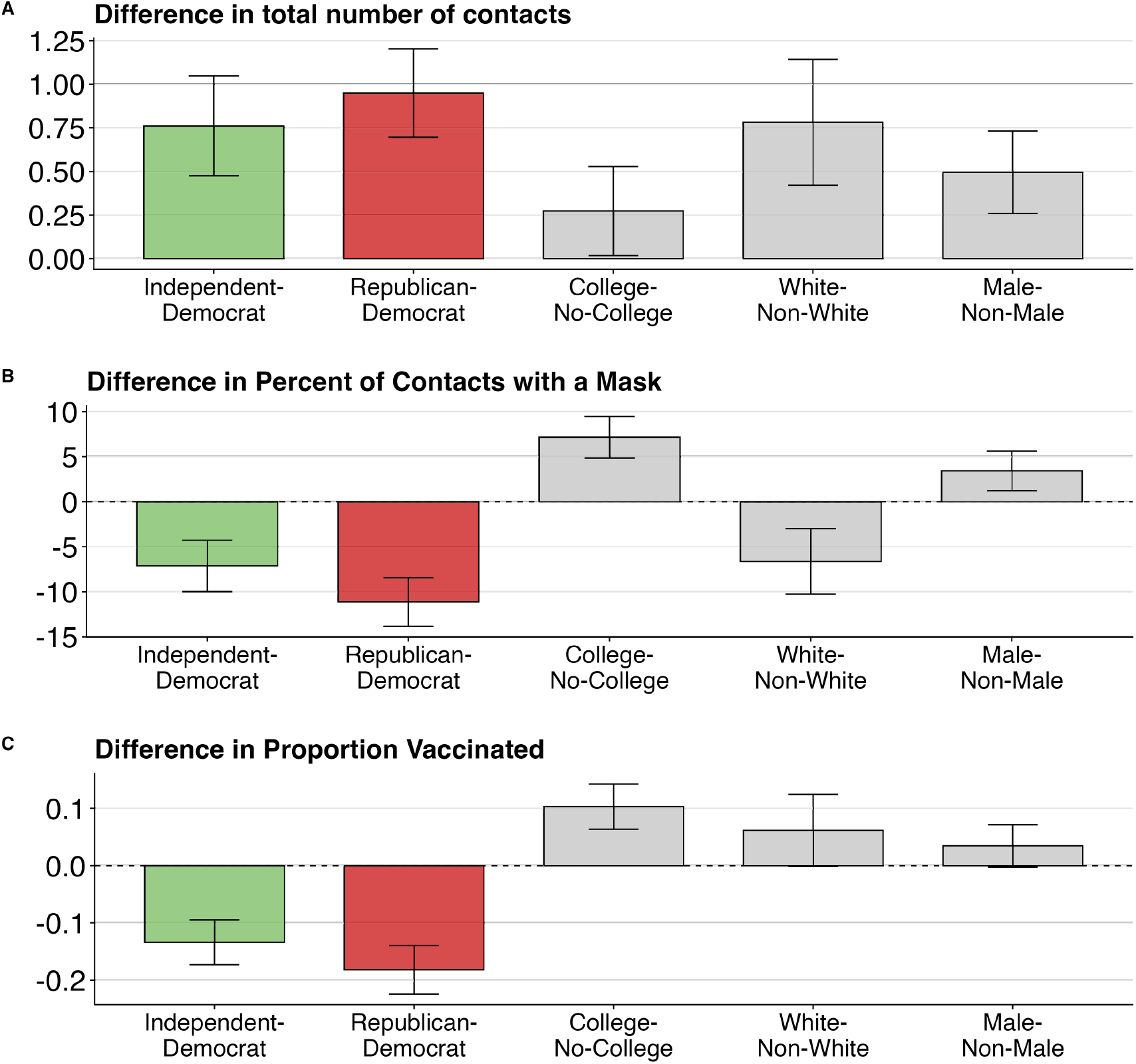
Partisan and Demographic Group Differences in Health Behaviors Among Adults Age 65 and Older. Comparison of partisan and demographic group differences for three health behaviors: (A) total reported in-person contacts from the previous day; (B) percentage of contacts involving mask use among those reporting previous-day contacts, based on detailed information from three selected contacts per respondent; and (C) estimated vaccination rates reported in Wave 6 of BICS in May 2021. Effect sizes are coefficients from univariate linear regression models estimated separately for each demographic characteristic and behavior. Sample includes respondents age 65 and older across six BICS survey waves (April 2020 – May 2021), except Panel C. Error bars show 95% confidence intervals.

**Figure S3:**
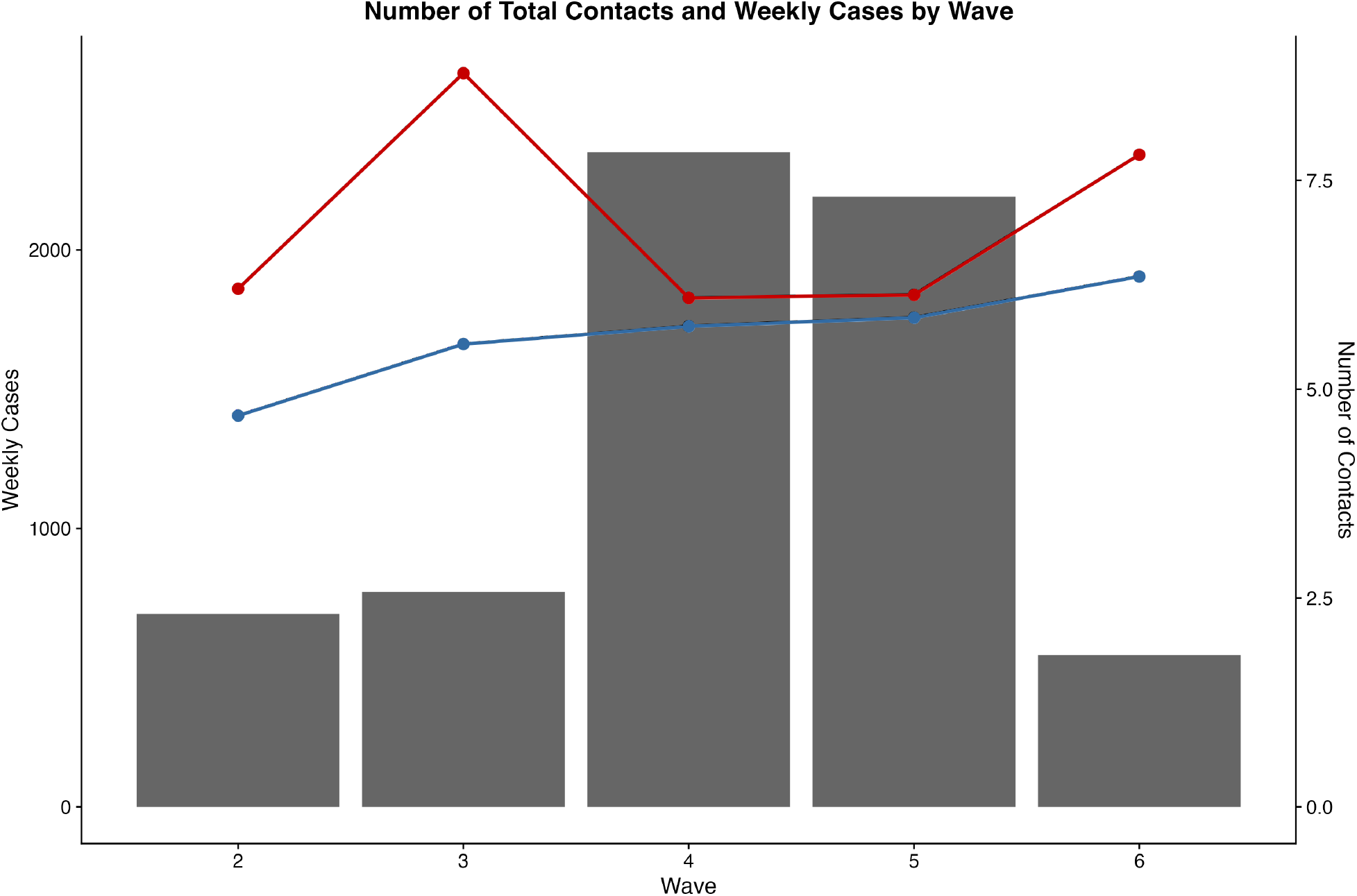
Partisan Contacts and National Weekly COVID-19 Cases Over Time. Average daily in-person contacts by partisan affiliation (left axis) shown alongside national weekly COVID-19 case counts (right axis, gray bars) across six BICS survey waves (April 2020 – May 2021). Republicans (red), Democrats (blue), and Independents (green) are shown with 95% confidence intervals. Contact rates for all partisan groups generally declined during periods of high case counts, though Republicans consistently maintained higher contact levels than Democrats throughout the study period.

**Figure S4:**
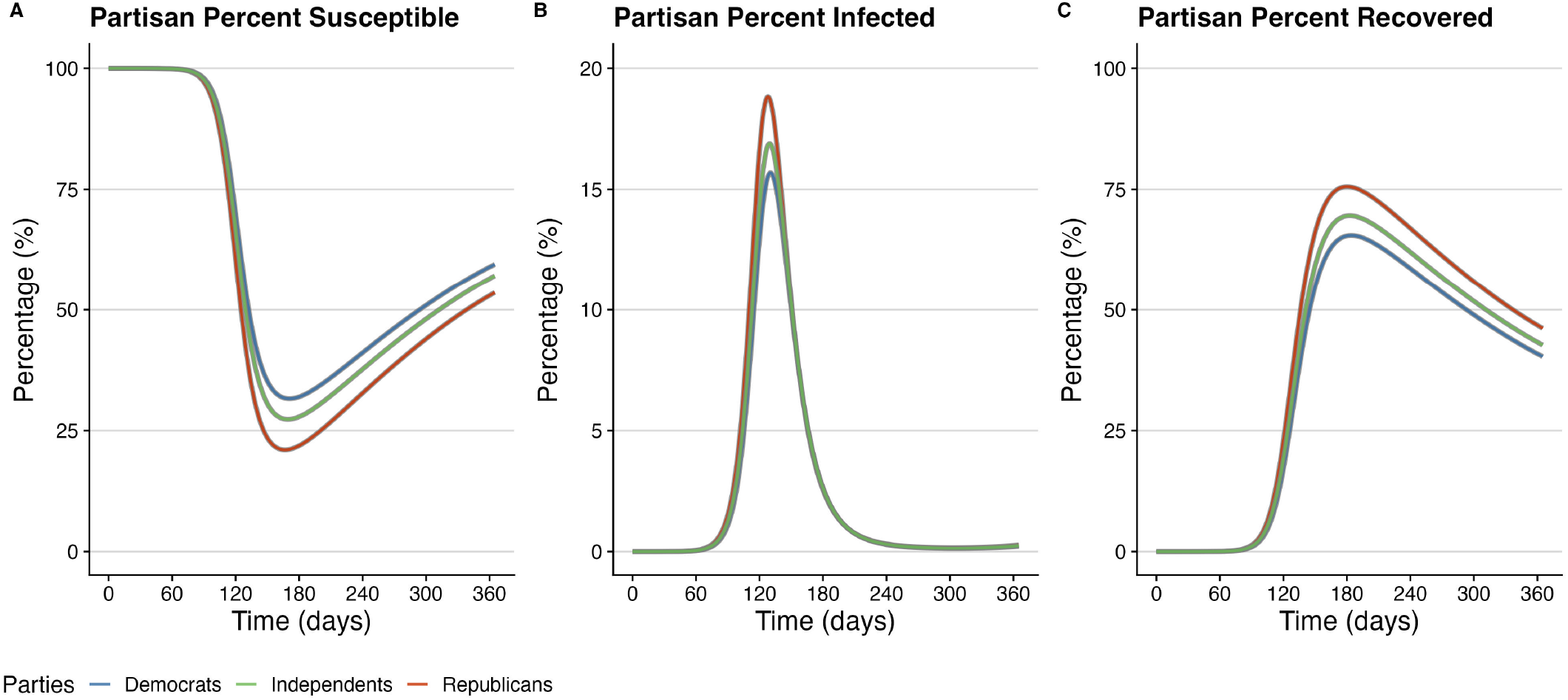
Multi-Panel Epidemic Outcomes Incorporating Partisan Behavioral Patterns Over Time. Results from a three-party SIR epidemiological model incorporating behavioral data pooled from all six BICS survey waves (April 2020 – May 2021), assuming random partisan mixing (*β* = 1) and an evenly split electorate over a one-year simulation. Panel (A) shows daily incidence rates per 10,000 for Republicans (red), Democrats (blue), and Independents (green). Panel (B) shows cumulative incidence over time. Panel (C) shows cumulative mortality. Panel (D) shows the percentage of each group currently infected over time. All model parameters sourced from published literature as detailed in Table S7.

**Figure S5:**
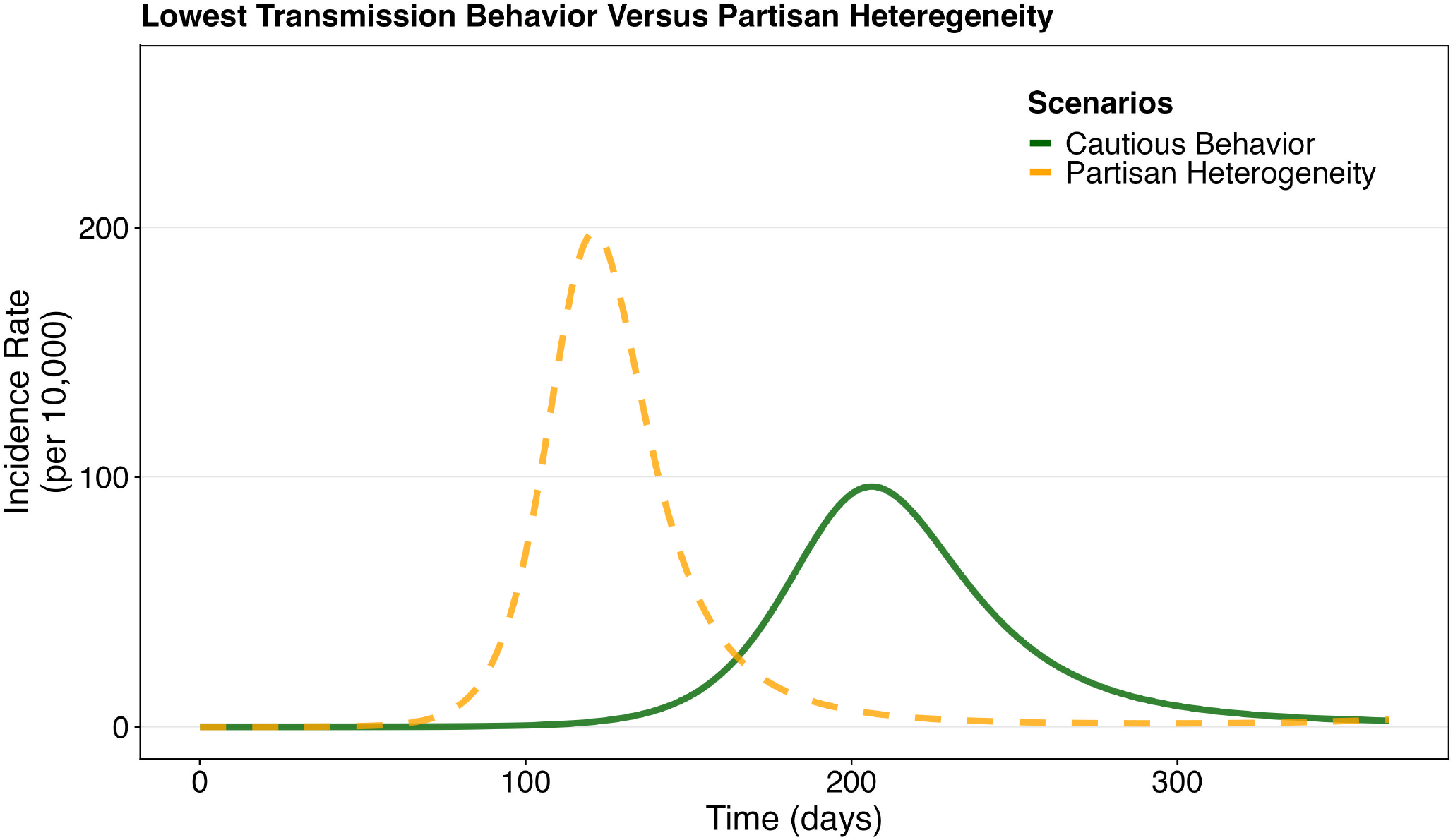
Sensitivity Analysis: Epidemic Outcomes Under Increased Protective Behavior Adoption. Results from a three-party SIR model with elevated background rates of protective behavior adoption (*π*_*i*_) compared to baseline estimates. This scenario examines how increased caution across all partisan groups affects epidemic dynamics, including incidence rates, cumulative infections, and mortality over a one-year simulation. All other model parameters remain as specified in Table S7.

**Figure S6:**
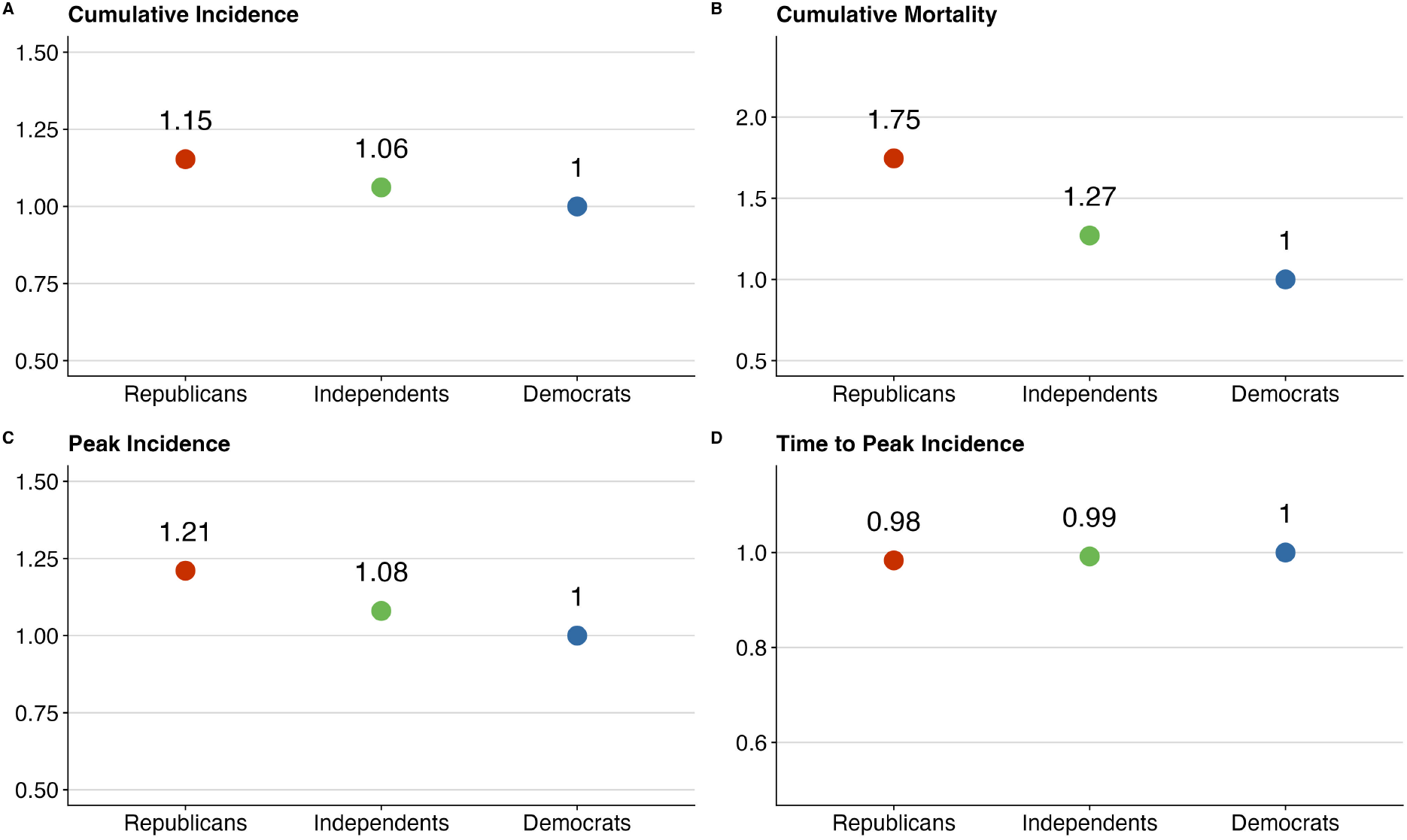
Results from a three-party SIR epidemiological model incorporating data pooled from all six BICS survey waves (April 2020-May 2021), assuming random partisan mixing and an evenly split electorate over a one-year simulation. The model incorporates differential mortality by assigning higher average infection mortality rates for older groups (Republicans) as outlined in the methods, reflecting that a COVID-like disease would be deadlier for older people. Data points are shown relative to the reference group (Democrats), with Democrats always equal to 1. Values greater than 1 indicate numbers larger than Democrats whereas values less than 1 indicate numbers smaller relative to Democrats. Panel (A) displays relative cumulative incidence. Panel (B) displays the relative cumulative mortality. Panel (C) displays the relative peak percentage of each group infected. Panel (D) displays the relative time to reach peak infection. All model parameters sourced from published literature as detailed in Table S7.

**Figure S7:**
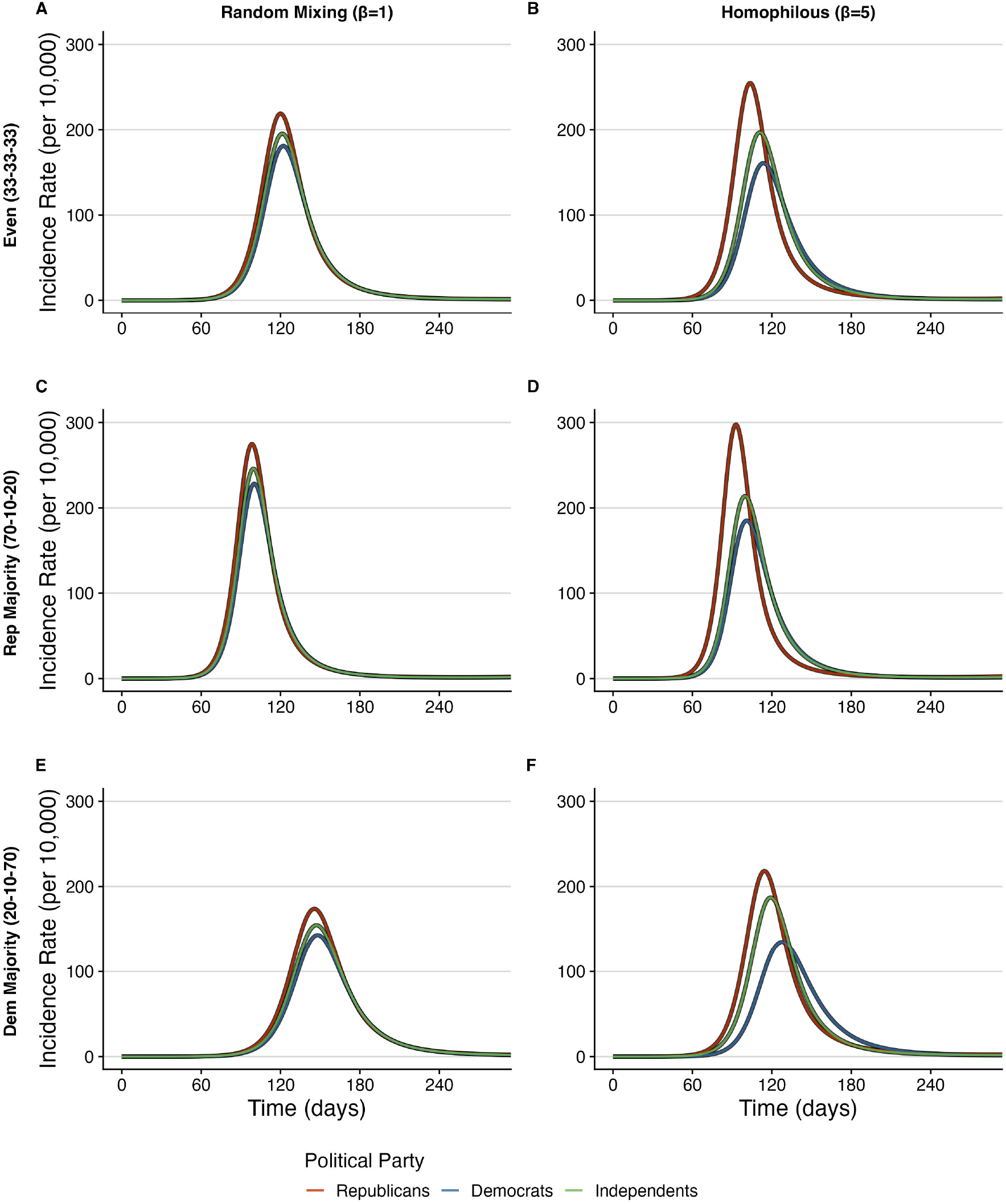
Partisan-specific incidence rates from SIR epidemiological models incorporating data from all six BICS survey waves (April 2020-May 2021). Left column: random mixing (*β* = 1); right column: homophilous mixing (*β* = 5). Rows show different population compositions: (A-B) even electorate (33-33-33), (C-D) Republican majority (70-10-20), (E-F) Democrat majority (20-10-70). Republican (red), Democrat (blue), and Independent (green) incidence rates per 10,000 shown over one year. Partisan disparities in infection rates increase with both Republican population share and homophily.

**Figure S8:**
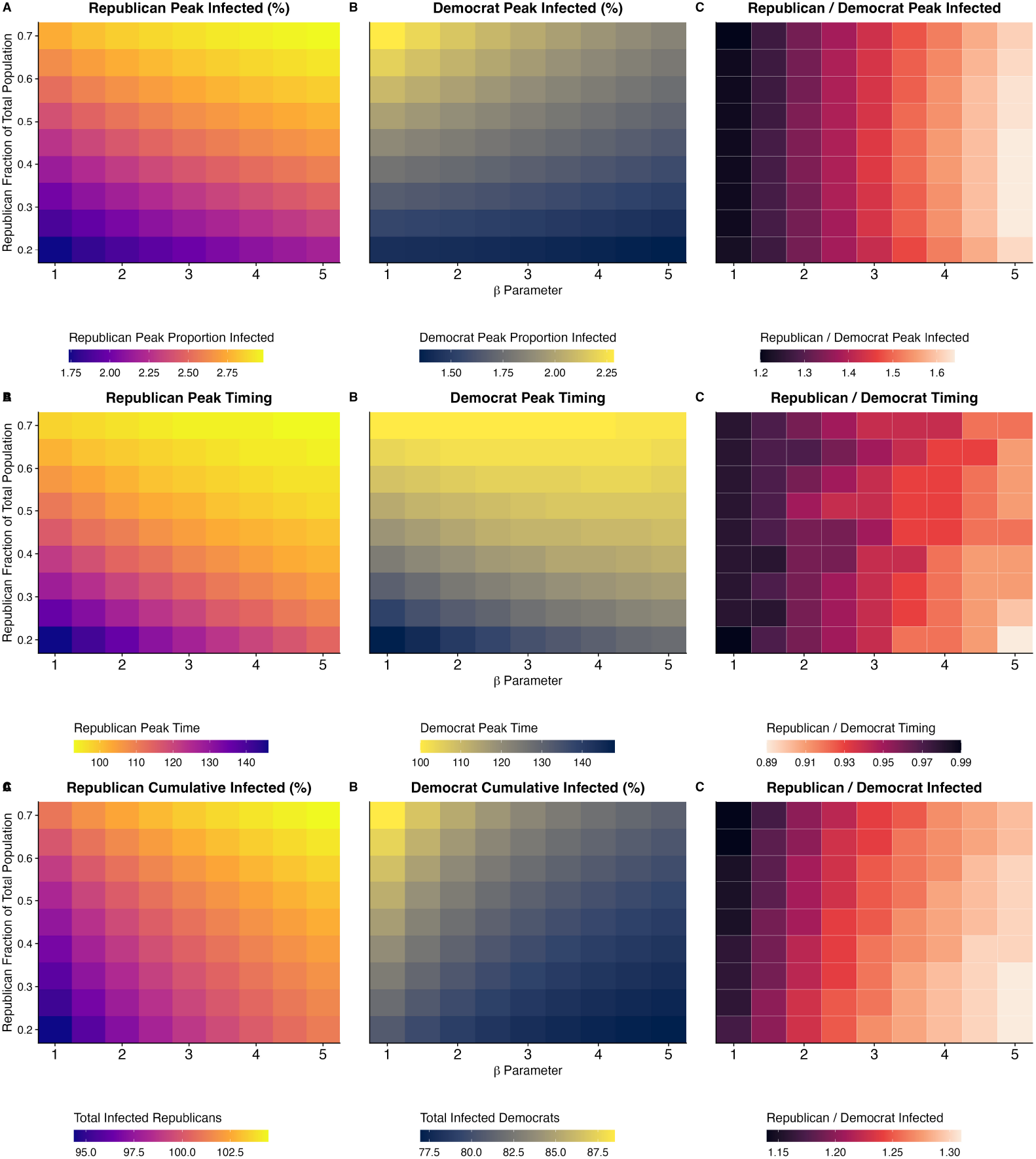
The effect of varying partisan homophily and population composition on peak infected. Results from sensitivity analysis varying Republican homophily parameters (*β*: 1.0-5.0, 51 steps) and population fractions (Republican: 20-70%, Democrat: 70-20%). Each simulation uses the three-party SIR model with base parameters from Table S7 to examine epidemic outcomes including peak infection rates, cumulative incidence, and mortality by partisan group.

**Figure S9:**
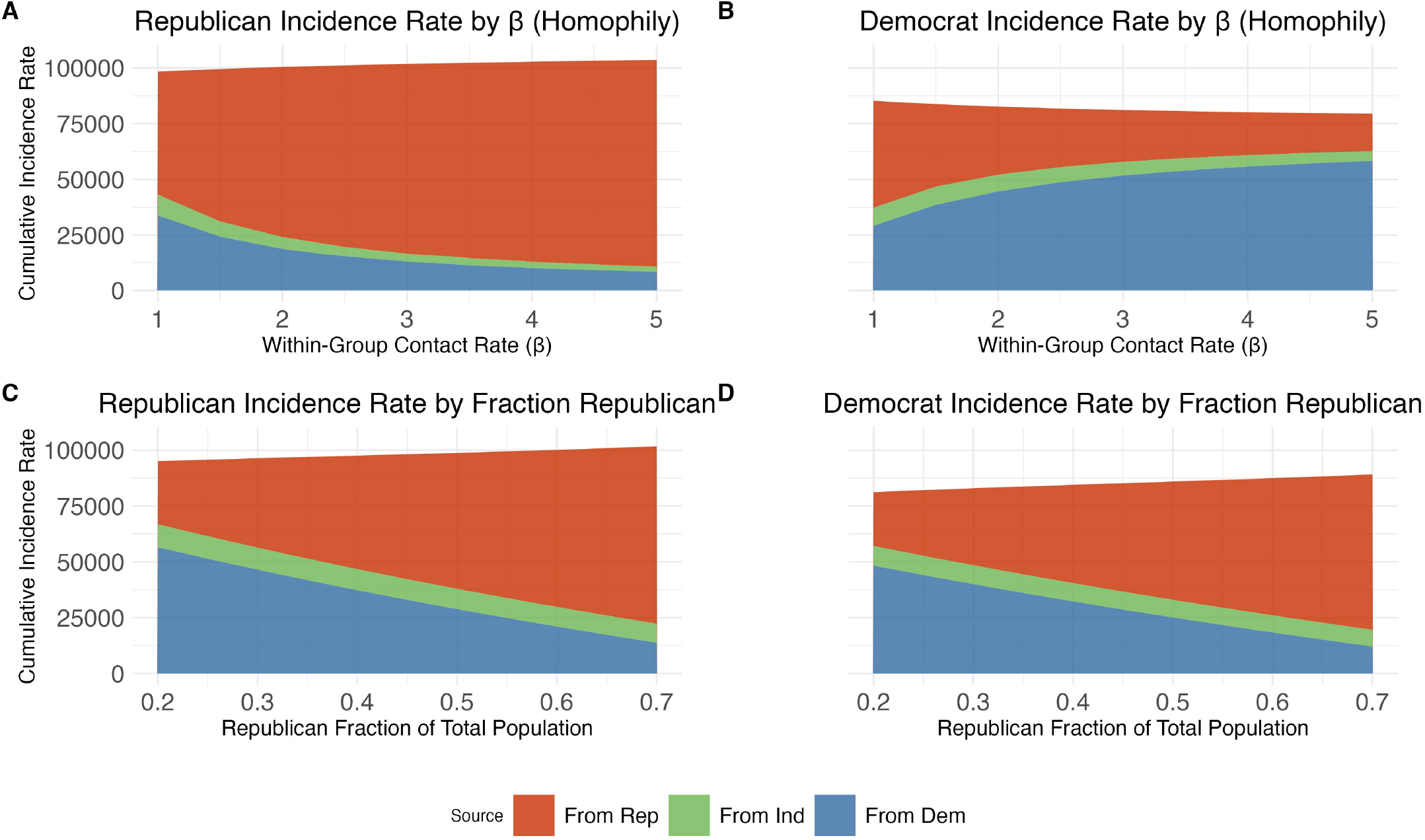
The effect of varying partisan homophily and population composition on outbreak size. Results from sensitivity analysis varying Republican homophily parameters (*β*: 1.0-5.0, 51 steps) and population fractions (Republican: 20-70%, Democrat: 70-20%). Each simulation uses the three-party SIR model with base parameters from Table S7 to examine outbreak size by partisan group. Panel (A) shows Republican outbreak size as a function of homophily (*β*), (B) shows Democrat outbreak size as a function of homophily (*β*), (C) shows Republican outbreak size as a function of Republican population fraction, and (D) shows Democrat outbreak size as a function of Republican population fraction.

**Figure S10:**
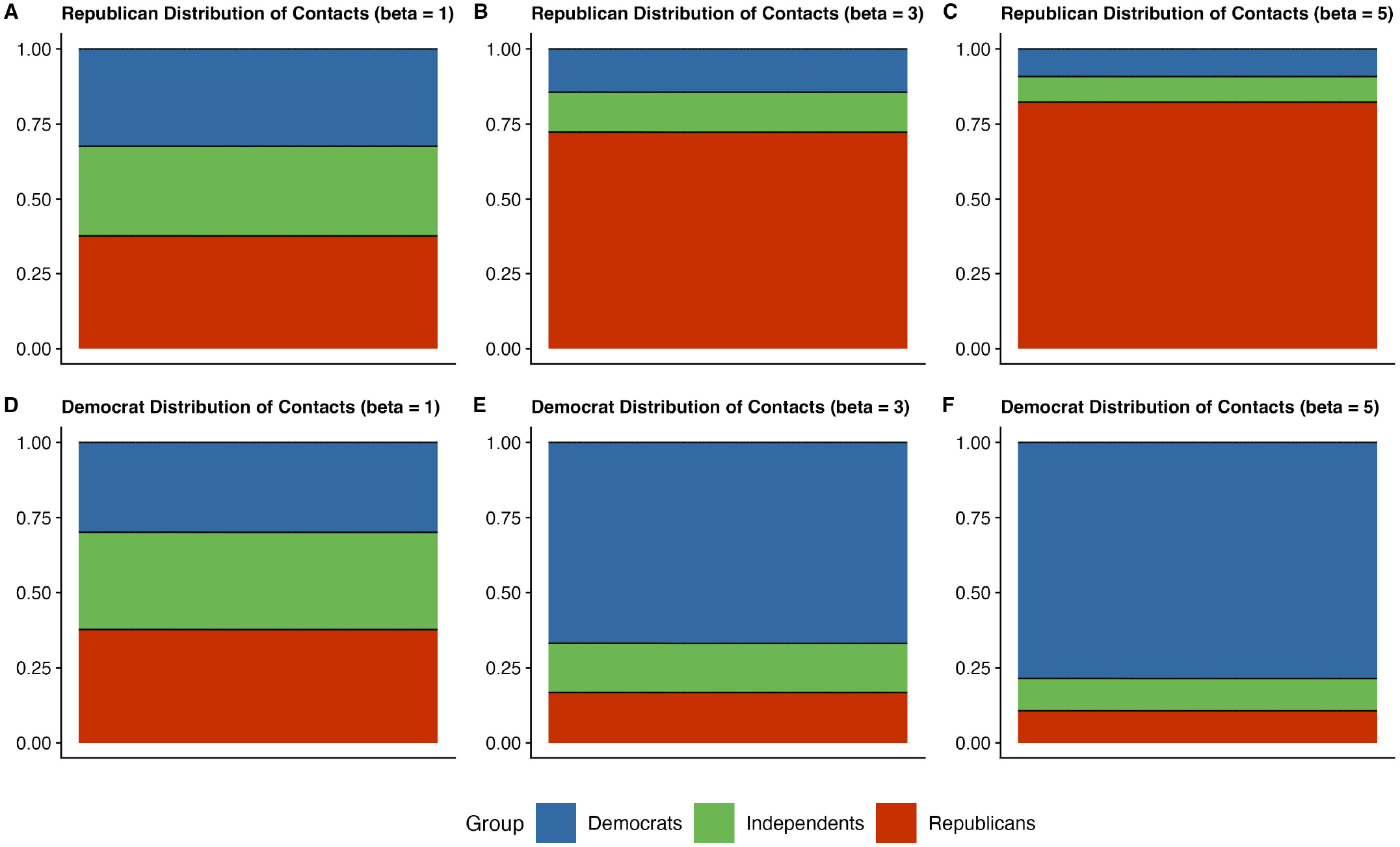
Share of contacts with other partisan groups based on homophily parameters (*β* = 1, 3, 5) in an evenly split population. Each stacked bar shows the proportion of contacts with Democrats (blue), Independents (green), and Republicans (red). Panels (A), (B), and (C) show Republican contact patterns under no homophily (*β*=1), moderate homophily (*β*=3), and high homophily (*β*=5), respectively. Panels (D), (E), and (F) show corresponding Democrat contact patterns under the same homophily conditions. As homophily increases, both Republicans and Democrats contact more members of their own group and fewer from opposing groups.

**Figure S11:**
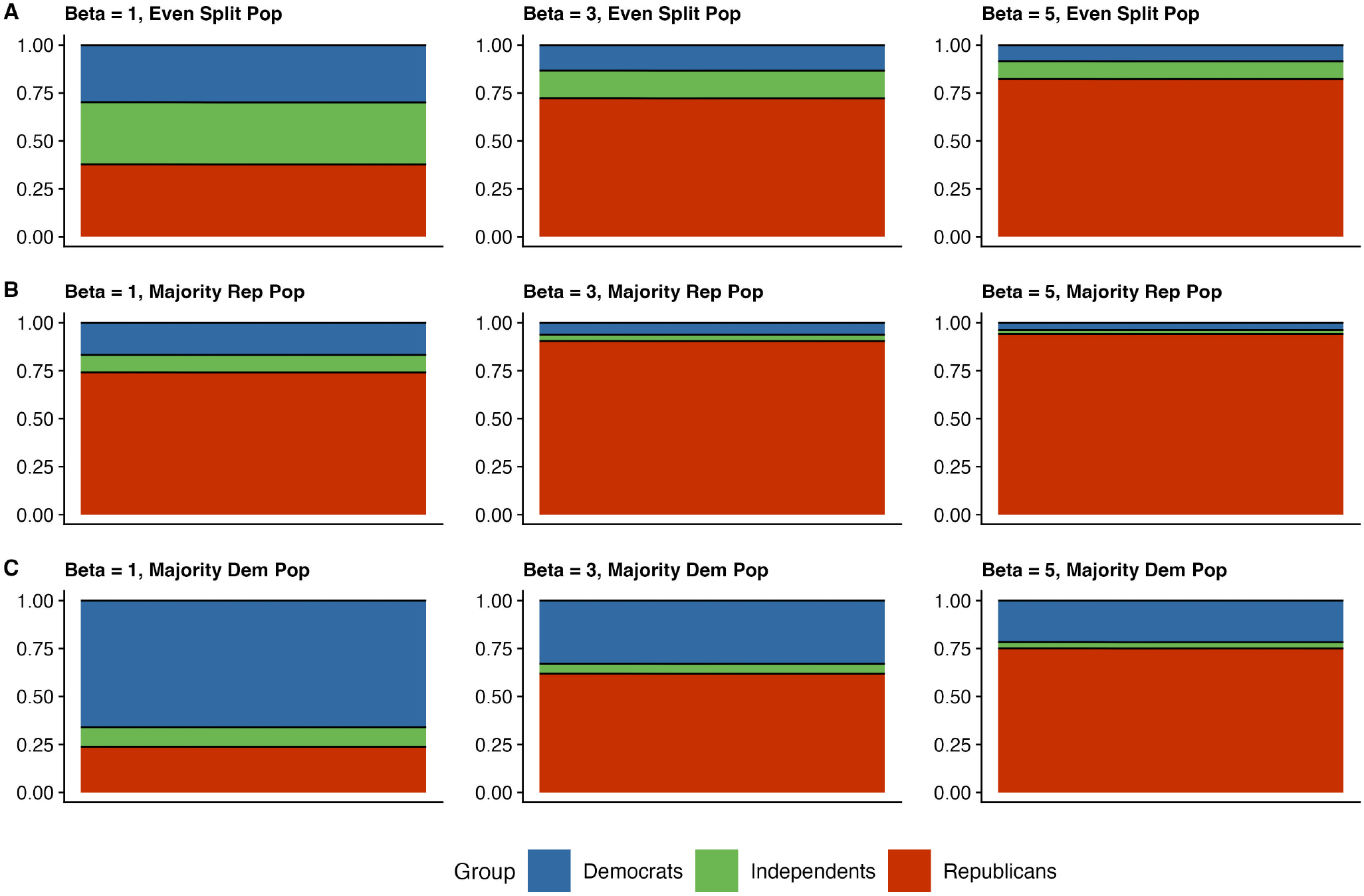
Republican Contact Patterns Under Varying Homophily and Population Composition. Share of Republican contacts with other partisan groups based on homophily parameters (*β* = 1, 3, 5). Each stacked bar shows the proportion of Republican contacts with Democrats (blue), Independents (green), and other Republicans (red). Row A displays results for an evenly split population, Row B for a Republican majority population, and Row C for a Democrat majority population. Columns show increasing homophily from left to right: no homophily (*β*=1), moderate homophily (*β*=3), and high homophily (*β*=5). Even without homophily, Republicans have more same-party contacts in Republican-majority populations due to baseline demographics. As homophily increases, Republicans increasingly contact members of their own group while reducing contacts with opposing partisan groups.

**Figure S12:**
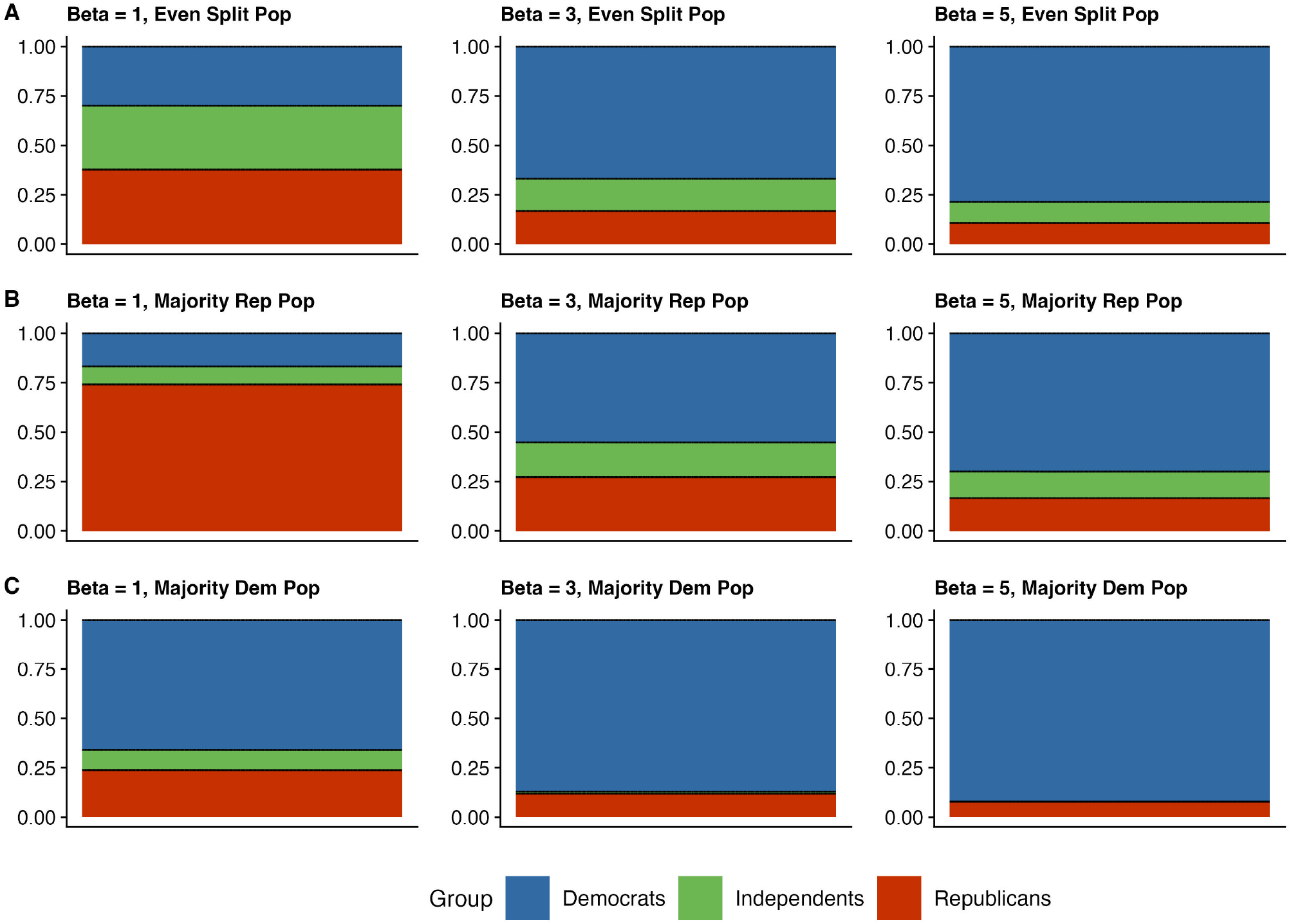
Democrat Contact Patterns Under Varying Homophily and Population Composition. Share of Democrat contacts with other partisan groups based on homophily parameters (*β* = 1, 3, 5). Each stacked bar shows the proportion of Democrat contacts with Democrats (blue), Independents (green), and Republicans (red). Row A displays results for an evenly split population, Row B for a Republican majority population, and Row C for a Democrat majority population. Columns show increasing homophily from left to right: no homophily (*β*=1), moderate homophily (*β*=3), and high homophily (*β*=5). Even without homophily, Democrats have more same-party contacts in Democrat-majority populations due to baseline demographics. As homophily increases, Democrats increasingly contact members of their own group while reducing contacts with opposing partisan groups.

